# An integrated single-cell atlas of checkpoint inhibitor-induced liver injury links shared liver-tumour CD8+ T cell clones to cytotoxicity and macrophage crosstalks

**DOI:** 10.64898/2026.04.27.26351449

**Authors:** Sarp Uzun, Simon Haefliger, Carl P. Zinner, Asmita Pant, Amke Beenen, Nikolai Bendik, Hélène Heusler, Anna K. Stalder, James Whipman, Kirsten D. Mertz, Jürg Vosbeck, Alfred Zippelius, Markus H. Heim, Natalie de Souza, Christine Bernsmeier, Heinz Läubli, Bernd Bodenmiller, Matthias S. Matter

## Abstract

Immune checkpoint inhibitors (ICIs) have transformed cancer therapy, but they can also induce immune-related adverse events (irAEs). Checkpoint inhibitor-induced liver injury (ChILI) is among the most frequent irAEs, yet its pathophysiology remains poorly understood. Here, we assembled a cohort of liver biopsies from cancer patients with ChILI and used a multi-modal analysis integrating single-cell spatial proteomics, bulk T cell receptor (TCR) sequencing and single-cell spatial transcriptomics to construct the first single-cell spatial atlas of ChILI. Integrating bulk and spatial TCR analyses revealed expanded T cell clones with a cytotoxic CD8+ phenotype that were shared between the liver and tumour. Intercellular communication analyses further indicated close interactions between the shared T cell clones and macrophages involving CCL5-CCR1 signalling. Our work provides in situ evidence of tumour-associated T cell contributions to ChILI. Furthermore, it establishes a framework for gaining mechanistic insights into ChILI and identifying therapeutic targets.

## INTRODUCTION

Immune checkpoint inhibitors (ICIs) have transformed cancer therapy. They provide substantial clinical benefits across a broad spectrum of advanced malignancies, such as metastatic melanoma, non-small cell lung cancer, and hepatocellular carcinoma.^1–3^ However, ICIs can also cause immune-mediated toxicities, collectively referred to as immune-related adverse events (irAEs), which can be severe and potentially treatment-limiting. These irAEs can affect various organs, most commonly the skin, colon, liver, lungs, and endocrine organs.^4,5^ As ICIs are now integral to the treatment of a growing number of malignancies, the clinical management of irAEs has become a central part of oncology care.

The liver is the third most frequently affected organ by ICIs. Liver irAE, also known as checkpoint inhibitor-induced liver injury (ChILI), typically manifests within the first 1-3 months of ICI-therapy.^6,7^ Despite its clinical relevance, little is known about the mechanisms underlying ChILI. Cytotoxic CD8+ T cells appear to be central mediators of ChILI by producing granzymes, IFN-γ and TNF-α.^8–10^ An important role has also been attributed to CD4+ T cells^8^ and myeloid cell subpopulations, such as CCR2+ CD163+ monocyte-derived macrophages.^9,11^ It is also thought that epitope sharing is one of the main mechanisms responsible for irAE, in which tumour neoantigens lead to activation of T and B cells that target similar self-antigens of healthy tissue.^12–14^ Although this mechanism has been implicated in other irAEs, it has not yet been demonstrated in the liver.^15^

Many insights into ChILI pathogenesis stem from *in vitro* or murine studies. However, there is a general lack of systematic, tissue-based characterisation of the human liver immune microenvironment in ChILI. Furthermore, the contribution of T cell clones shared between the tumour and liver, as well as the origin and signalling pathways underlying the cytotoxic milieu in ChILI, remain unexplored. Therefore, in this study, we assembled a cohort of human liver biopsies with ChILI to characterise the immune infiltrate comprehensively and explore the contribution of T cell clones shared between the liver and tumour. By integrating single-cell spatial proteomics, bulk TCR sequencing and single-cell spatial transcriptomics, we have elucidated the processes underlying ChILI and identified potential therapeutic targets.

## RESULTS

### Design of the ChILI and control cohort

We built a cohort of formalin-fixed and paraffin-embedded (FFPE) liver biopsy specimens from 35 cancer patients who developed ChILI under ICI therapy (**Fig. 1A**, **Supplementary Table 1-2**). The mean age of patients was 62.6 years and 47% were female. The patients were treated with ICI for different types of cancers, mainly melanoma, lung and renal cancer. Half of the patients (46%) had a combination therapy, mostly with ipilimumab and nivolumab. The remaining patients received monotherapy, mainly with pembrolizumab. All patients had elevated liver enzyme levels, and most patients (80%) had grade 3 or 4 ChILI, i.e. severe or life-threatening, according to the Common Terminology Criteria for Adverse Events (CTCAE; version 5.0). Based on liver enzyme levels^16^, 66% (n = 23) were classified as ChILI hepatitis, 20% (n = 7) as ChILI cholangitis, and 14% (n = 5) as mixed forms (**Supplementary Table 2**). Because liver biopsies are not performed in ICI-treated patients who do not develop ChILI, we instead used an external control group. This comprised 16 cancer patients who had been newly diagnosed with a liver metastasis and had not undergone any cancer therapy. They underwent a biopsy of non-tumoural liver tissue, which showed histologically uninflamed tissue with normal liver enzyme levels. There was no difference in age or gender between the ChILI and the control cohort (**Fig. 1A, Supplementary Table 2**). For our study, we used various spatial and single-cell techniques for detailed analysis of immune cell populations in these liver biopsy samples. Not all assays were performed on each patient sample, mainly due to limited availability or quality of tissue (**Fig. 1B, Supplementary Table 1**).

**Fig. 1.**
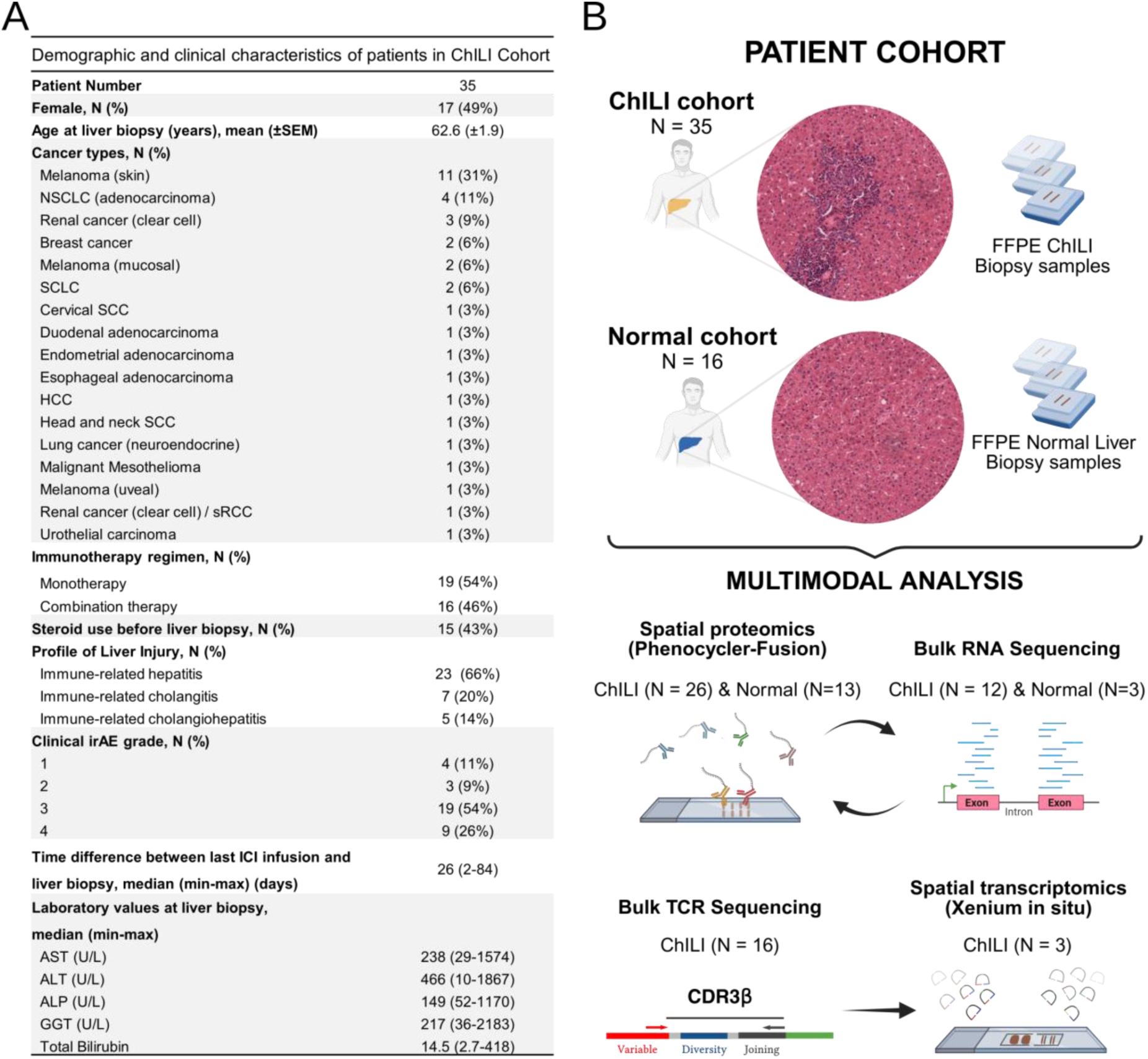
Demographic and clinical characteristics of ChILI cohort and experimental design of the study. (A) Demographic and clinical characteristics of patients included in the ChILI cohort. (B) Schematic overview of the experimental workflow for multimodal analysis of liver biopsies, integrating high-dimensional spatial proteomics and bulk RNA-sequencing and TCR sequencing with spatial transcriptomics.

### A highly multiplexed single-cell map of immune cells in ChILI

To characterise the immune microenvironment in ChILI, we performed multiplex immunofluorescence (mIF) with the Phenocycler-Fusion (PCF) system. Samples were imaged using a 40-marker antibody panel (**Fig. 2A, Extended Data Fig. 1, Supplementary Table 3**), which enabled the phenotypic characterisation of the main immune cell types, but also parenchymal and non-parenchymal cells of the liver. A total of 556’897 single cells were profiled across 39 liver biopsies from 26 ChILI and 13 normal control patients. Image processing and single-cell segmentation were performed using an established image analysis workflow^17,18^ (**Fig. 2B-C**). No batch effect across patients or cohorts was found (**Extended Data Fig. 2A-D**). Each segmented cell was mapped to its corresponding liver anatomical compartment - portal, lobular, or interface - using compartment masks, and the accuracy of these assignments was validated by visual inspection (**Extended Data Fig. 2E)**. To define major cell phenotypes, we applied Louvain clustering^19^ on the expression data of key lineage markers (see methods). We defined 12 distinct broad cell types: hepatocytes, CD4+ T cells, myeloid/macrophages, CD8+ T cells, stromal cells, plasma cells, vessels, NK cells, B cells, neutrophils, cholangiocytes, and eosinophils. These assigned single-cell phenotypes were visualized using a t-distributed stochastic neighbour embedding (t-SNE) **(Fig. 2D)**. As expected, samples with ChILI exhibited increased fraction in almost all immune cells compared to normal liver tissue (**Fig. 2E, Extended Data Fig. 3A-B)**. However, changes in cell density and cell fraction were significant only for CD8+ T cells and myeloid/macrophages (**Fig. 2F**), and - at much lower absolute levels - for plasma cells (**Extended Data Fig. 3A-B**).

**Fig. 2.**
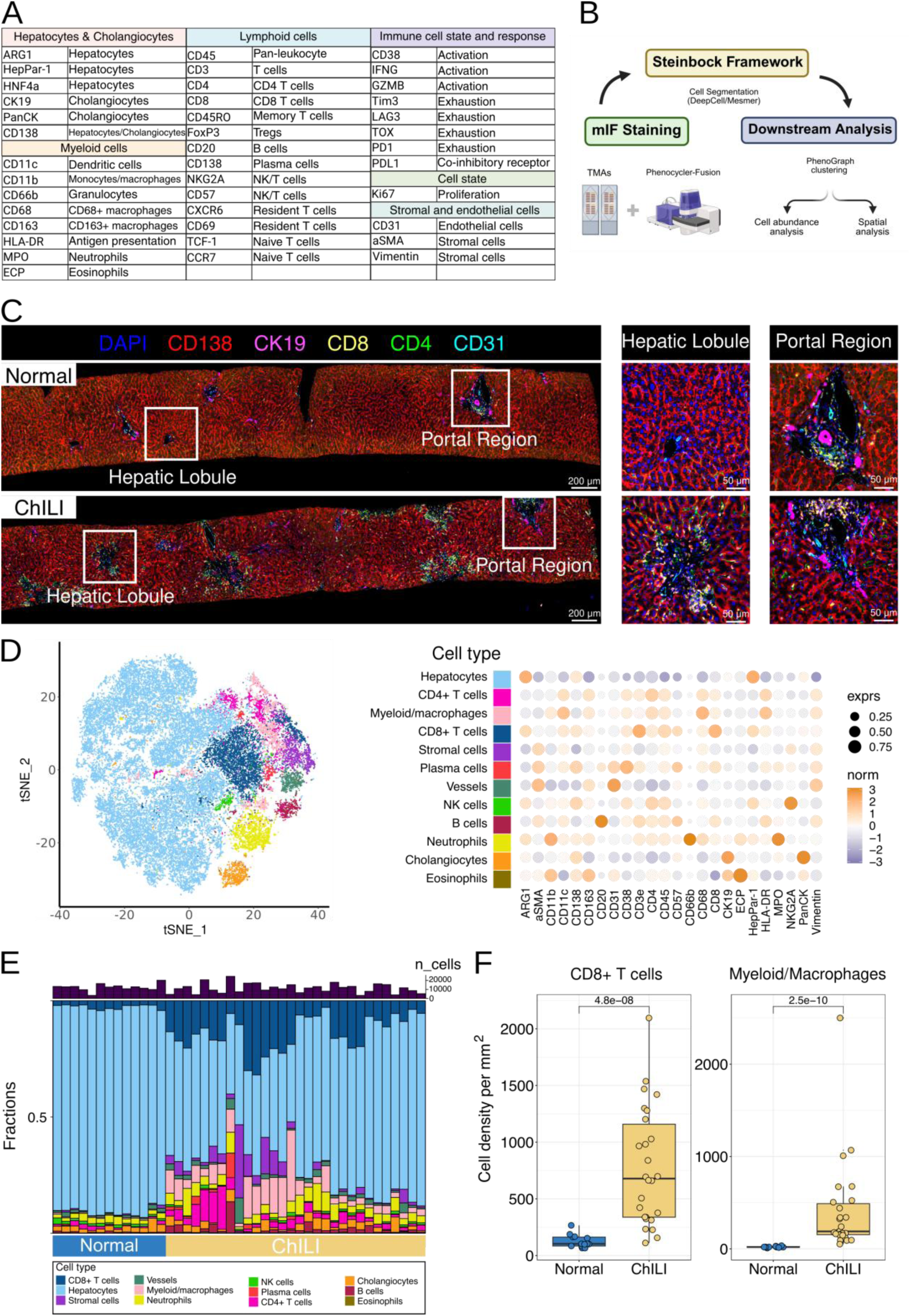
mIF imaging generates a single-cell map of ChILI. (A) Markers used for cell phenotyping, including lineage-defining and functional markers. (B) Scheme of the imaging analysis pipeline. Multiplexed images were generated, followed by cell segmentation, phenotyping, and assignment of cells to major cell types. (C) Representative liver biopsy image from normal (upper panel) and ChILI liver biopsy (lower panel) showing spatial distribution of major compartments, with insets highlighting hepatic lobule with central vein and portal region. Immunofluorescent markers include DAPI (nuclei), CD138 (hepatocytes), CK19 (biliary cells), CD8 and CD4 (T cells), and CD31 (vessels). (D) Left: t-distributed stochastic neighbour embedding (t-SNE) representation of single cells, coloured by assigned cell type. Right: Dot plot summarizing marker expressions across phenotypically defined cell types. Dot size indicates the fraction of cells expressing the marker, while colour intensity reflects scaled mean expression. Cell types are annotated along the y-axis and markers on the x-axis. (E) Barplot showing the fraction of each cell type per biopsy, stratified by patient and cohort (normal and ChILI). Colour coding corresponds to cell type, and vertical bars are grouped by disease condition and patient ID. (F) Boxplots show densities (cells/mm²) of CD8+ T cells and myeloid/macrophages across cohorts (normal vs ChILI). Statistical comparisons were performed using Wilcoxon rank-sum test with Benjamini-Hochberg correction; adjusted p-values are indicated.

### Spatially organized niches of CD8+ cytotoxic T cells and macrophages in ChILI

To dissect the immune microenvironment of ChILI in more detail, we applied Louvain clustering^19^ to CD8+ and CD4+ T cells as well as to myeloid/macrophages, using lineage- and state-defining markers (see Methods). We identified three CD8+ T cell subtypes: CD8+ cytotoxic T cells, CD8+ naive/central memory (CM) T cells, and CD8+ T cells without a specific marker profile based on our antibody panel and referred to as mixed CD8+ T cells (**Fig. 3A-B)**. We also characterised three CD4+ T cell subtypes: Tregs, TCF1+ CD4+ T cells, and CD4+ T cells without a specific marker profile referred to as CD4+ mixed T cells. Finally, we identified three myeloid/macrophage subtypes: CD68+ CD163^Hi^, CD68+ CD163^Lo^ and dendritic cells (**Fig. 3A-B)**. These immune subtypes were then visualized using a t-SNE (**Fig. 3B)**. ChILI showed significantly higher cell density and concordantly increased cell fractions for all these immune cell subtypes, except for CD8+ naive/CM T cells and CD4+ mixed T cells, when compared to normal liver tissue (**Fig. 3C, Extended Data Fig. 4A**). The highest increase in absolute cell density was observed for CD8+ cytotoxic T cells, followed by CD8+ mixed T cells, CD68+ CD163^Hi^ and CD68+ CD163^Lo^ macrophages (**Fig. 3D, Extended Data Fig. 4A-B).** To confirm these observations obtained by mIF, we analysed the transcriptomes of a subset of 12 ChILI and 3 control liver biopsies using bulk RNA (**Extended Data Fig. 5A**). Dimension reduction analysis clearly separated ChILI from normal liver (**Extended Data Fig. 5B-C**). Differential gene expression and gene set enrichment analysis (GSEA) revealed robust immune activation in ChILI, including the enrichment of the IFN-γ and TNF-α pathways, whereas pathways involved in metabolism were downregulated (**Extended Data Fig. 5D-E).** Immune deconvolution using MCP-counter^20^ revealed a significant increase in T cells, CD8+ T cells, and cytotoxicity score (**Extended Data Fig. 5F-G**). Additionally, there was an increase in myeloid dendritic cells, macrophages/monocytes, and NK cells. In contrast, B cells and neutrophils were not significantly altered. Therefore, bulk transcriptomics confirmed the increase of CD8+ T cells, CD8+ cytotoxic T cells, and myeloid/macrophages in ChILI as previously found by mIF.

**Fig. 3.**
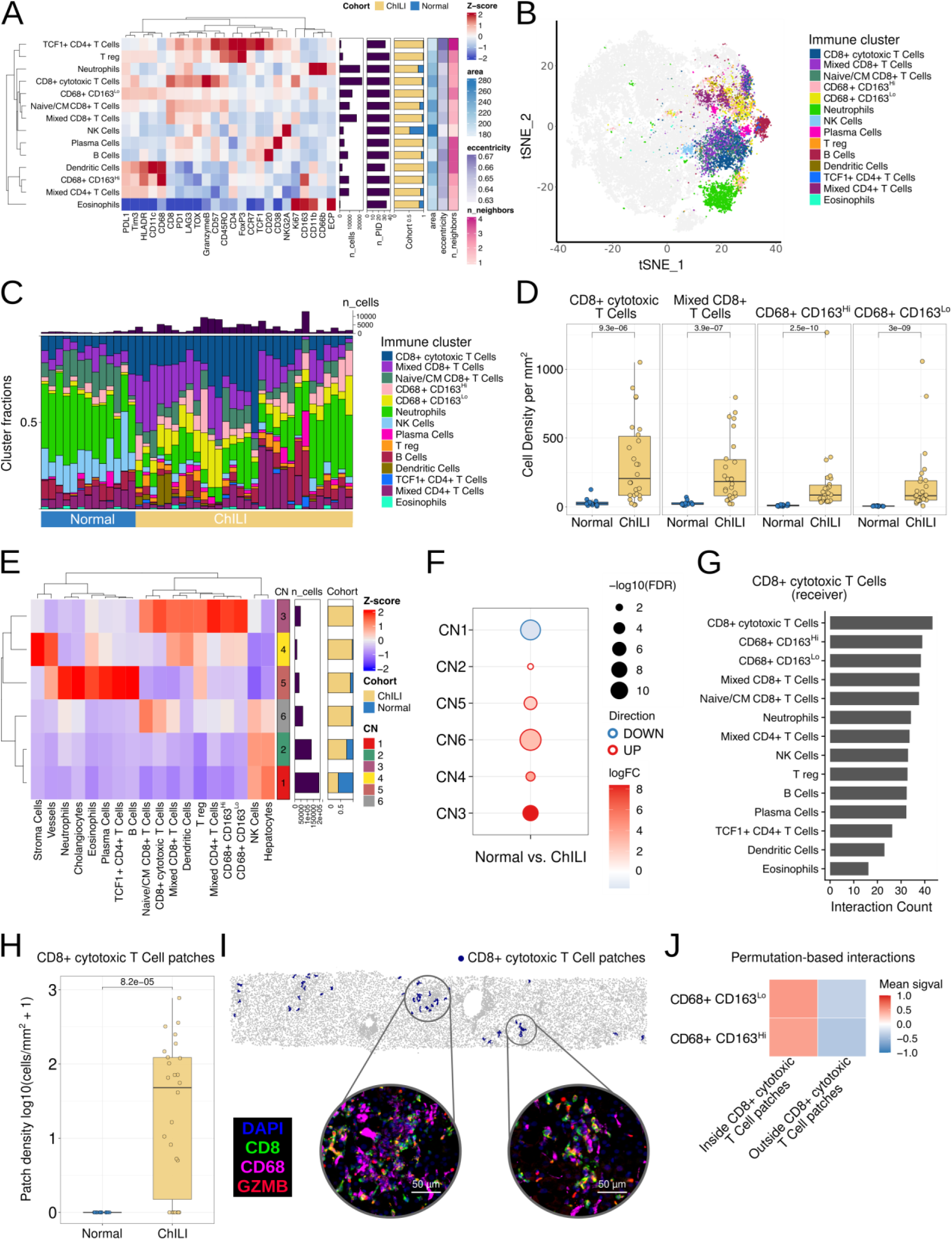
Compartment analysis reveals spatially distinct, organized immune niches in ChILI. (A) Heatmap of expression (Z-score) per immune cell subtypes. The plots (right) show the total numbers of cells and patients per immune subtypes alongside the fraction per cohort and average spatial single cell information (area, eccentricity, number of neighbours). Rows and columns are hierarchically clustered using Ward.D2. All markers indicated were used for graph-based clustering. (B) tSNE visualization of all immune cells from ChILI and normal liver samples, coloured by immune cell subtype identity. (C) Stacked barplot depicting the fraction of each immune cell subtype per biopsy, stratified by sample and cohort. (D) Boxplots of the indicated immune cell subtypes densities (cells/mm²) across cohorts (normal vs ChILI). Statistical comparisons were performed using Wilcoxon rank-sum test with Benjamini-Hochberg correction; adjusted p-values are indicated. (E) Hierarchical clustering heatmap of cellular neighbourhoods (CN1–CN6) defined from immune cell subtypes. Columns are immune cell subtypes; rows are CNs. Values are z-scored immune cell subtype frequencies within each CN (red = enriched, blue = depleted). Annotations (right) show: cohort label (ChILI vs normal), the CN identifier, the number of cells contributing to each CN (n_cells), and cohort composition of CNs. (F) Differential abundance of CNs between normal and ChILI, shown as a bubble plot. Each dot is one CN; colour encodes log fold-change (red = higher in ChILI, blue = lower in ChILI relative to normal), size encodes significance (–log10 FDR). CNs with larger red bubbles are expanded in ChILI; blue bubbles indicate depletion. (G) Cell–cell interaction profile for CD8+ cytotoxic T cells as receiver cell. Bars show the absolute interaction count with each indicated partner, ranked from most to least frequent. (H) Box–jitter plot comparing the CD8+ cytotoxic T cell patch density per sample between normal and ChILI cohorts. The y-axis shows log10 (cells/mm² + 1). For each sample, density was computed as the number of cells assigned to CD8+ cytotoxic T cell patches divided by the tissue area. Boxes indicate median and IQR (whiskers = 1.5×IQR); points are individual samples; two-sided Wilcoxon test was used to compare cohorts. (I) Representative liver biopsy (ChILI_33) illustrating CD8+ cytotoxic T cell patches (blue markers) over the full section (grey). Insets (scale bars = 50 μm) show high-magnification views of mIF staining in CD8+ cytotoxic T cell patches. DAPI (blue), CD8 (green), CD68 (magenta) and GZMB (red). (J) Permutation-based interaction enrichment analysis in ChILI showing spatial interactions between macrophages (CD68+CD163^Hi^ and CD68+CD163^Lo^) and CD8+ cytotoxic T cells located within (patch+) or outside (patch−) CD8+ cytotoxic T cell patches. Heatmap values represent the mean interaction significance score (sigval) across samples, with positive values indicating enriched interactions and negative values indicating depleted interactions.

At the time of liver biopsy, about half of the ChILI patients received immunosuppressive therapy. Although most patients received treatment for only a few days, and all had elevated liver enzymes, we wanted to assess whether immunosuppressive treatment altered the composition of immune cells. T-SNE embedding and compositional analysis (**Extended Data Fig. 6A-B**) revealed no separation between treated (n = 14) and untreated (n = 12) patients. Similarly, there were no significant differences in immune cell types between treated and untreated patients, except that treated patients showed a trend towards reduced CD68+CD163Lo macrophages and a significant reduction in plasma cells (**Extended Data Fig. 6C**).

Next, we conducted spatial analyses to examine how immune cells are arranged within ChILI. First, we performed a cellular neighbourhood (CN) analysis, which enabled the identification of tissue regions with characteristic local composition of certain cell types.^21^ This analysis identified six distinct CNs (**Fig. 3E**). Differential abundance analysis between ChILI and normal samples revealed the highest fold changes for CN3 (**Fig. 3F**). Interestingly, CN3 is enriched in CD8+ cytotoxic T cells and myeloid/macrophages (CD68+CD163^Hi^, CD68+CD163^Lo^, dendritic cells) (**Fig. 3E**). In addition, we assessed cell-to-cell interaction counts between immune cell subtypes using the method by Shapiro^22^. This showed that, within the ChILI cohort, CD8+ cytotoxic T cells were preferentially adjacent to other CD8+ cytotoxic T cells, CD68+CD163^Hi^, and CD68+CD163^Lo^ macrophages (**Fig. 3G**), consistent with potential crosstalk between these populations. Interaction counts were calculated only within ChILI samples, because the normal liver cohort contained few immune cells, preventing a reliable comparative analysis. Finally, we explored the spatial aggregation of immune cell subtypes using the patch-detection framework by Hoch^23^, defining patches as spatially connected sets of at least five contiguous cells. Compared to all other immune subtypes, the highest patch density differential relative to normal liver was found for CD8+ cytotoxic T cells (**Extended Data Fig. 7A**). CD8+ cytotoxic T cell patches were absent in normal liver (**Fig. 3H)** and detected in 20 out of 26 ChILI patients. Furthermore, visual inspection showed that CD8+ cytotoxic T cell patches frequently contained macrophages (**Fig. 3I**). Indeed, cell permutation-based interaction testing revealed that CD8+ cytotoxic T cells within patches interacted more with macrophages (CD68+CD163^Hi^, and CD68+CD163^Lo^) than those located outside patches (**Fig. 3J, Extended Data Fig. 7B-C)**.

In conclusion, our proteomic and transcriptomic analyses indicate that ChILI is characterised by a predominance of CD8+ cytotoxic T cells and macrophages, which show evidence of spatial organization into local neighbourhoods and mutual interaction.

### T cell clones are shared between ChILI liver biopsies and matched tumour samples

T cell clone sharing between tumour and affected tissues has been documented in several irAEs, including myocarditis, pneumonitis and dermatitis, but not in ChILI.^12–14,24^. Therefore, we used TCR sequencing on isolated RNA to ask whether TCR sequences are shared between T cells of the liver and the tumour. The availability and quality of samples allowed for TCR repertoire analysis from 16 ChILI liver biopsies and their matched tumour samples (primary or metastatic). Shared T-cell clones were defined as having identical amino acid sequences in the complementarity determining region 3 (CDR3) of the TCR-β chain with the same variable (V) and joining (J) gene segments. Strikingly, every patient harboured shared T cell clones between the liver and tumour samples (**Fig. 4A, Extended Data Fig. 8, Supplementary Table 4**), with 6 to 190 shared clones per patient, accounting for 0.4%-2.58% of the combined TCR repertoire of liver and tumour samples from each patient (**Fig. 4A**). Notably, shared T cell clones had significantly higher clonality than tissue-unique clones, meaning, they were dominated by a few highly expanded clones (**Fig. 4B**). This indicated that shared T cell clones expanded within both the liver and the matched tumour and may contribute to anti-tumoural immune response as well as liver damage. To confirm that shared T cell clones do not only circulate within the blood but can traffic into the tissue, we designed BaseScope RNA in situ hybridization (RISH) probes recognizing the TCR-β CDR3 and flanking nucleotide mRNA sequences of two patients (P13: CASSPLTGELFF and P26: CATEGSLEQYF) (**Extended Data Fig. 9A, Supplementary Table 5**). RISH staining confirmed the presence of shared T cell clones in the liver (**Fig. 4D, Extended Data Fig. 9B**) and tumour tissue of both patients (**Fig. 4E, Extended Data Fig. 9C**). Furthermore, a combination of BaseScope RISH targeting the TCRβ CDR3 mRNA CASSPLTGELFF and immunohistochemistry for CD3 confirmed that TCR signals were localized to CD3+ T cells (**Fig. 4E**).

**Fig. 4.**
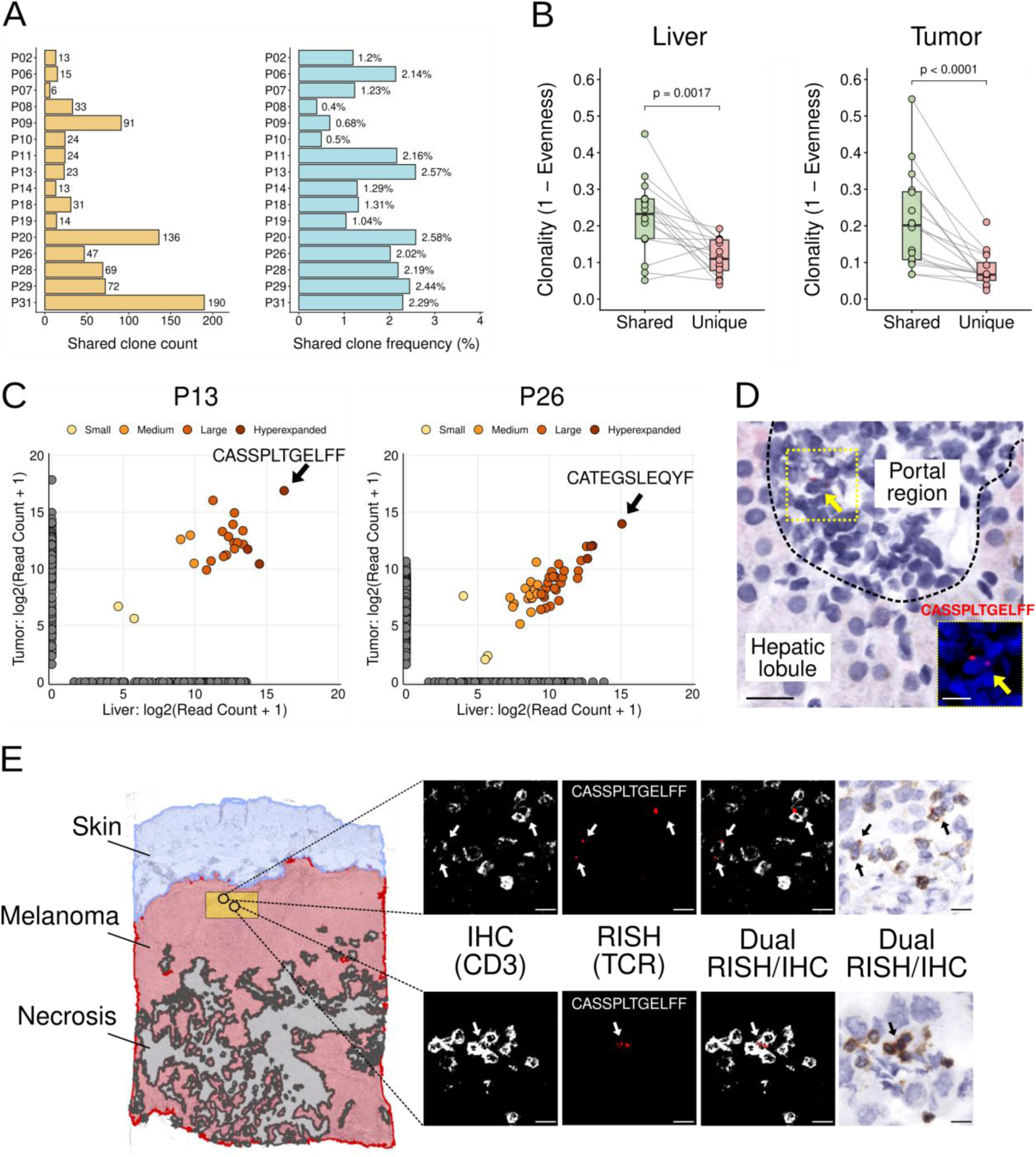
T cell immune repertoire of liver biopsy samples with ChILI and matched tumour samples. (A) Left: Absolute counts of shared T cell clones; right: Frequency of shared clones within the total TCR repertoire from liver and tumour samples of individual patients calculated with Jaccard similarity index. (B) Clonality of shared and non-shared clones in liver and tumour (Wilcoxon signed-rank test, p < 0.05). (C) Log2 normalised read counts of T cell clones from two patients (left: P13 and right: P26). Each dot indicates a clone. Dots from shared clones were coloured based on clonal abundance; dots from tissue-unique clones were coloured grey. (D) RNA in situ hybridization (RISH) using the indicated probe (CASSPLTGELFF) to identify the CDR3 mRNA sequence of the top shared hyperexpanded T cell clone in P13 liver biopsy sample. Scale bar = 10 µm. Inset shows the yellow rectangle having RISH+ cell. Blue: Hematoxylin. Red: CDR3 mRNA. (E) Dual RISH/IHC assay probing for RISH+ CD3+ cells. Left: Tissue masks were generated for skin, underlying tumour tissue (melanoma), and necrotic regions within the tumour of P13. The yellow area is magnified at right. IHC image shows CD3+ cells in white colour-deconvoluted image) or brown (brightfield image). RISH images show CD3+ cells positive for the indicated TCR sequence (red); RISH+CD3+ cells are shown by arrows. Scale bar = 10 µm.

In summary, TCR sequencing and BaseScope RISH revealed the presence of shared T cell clones in the livers and tumours of patients with ChILI.

### Shared T cell clones show a cytotoxic phenotype

Next, we analysed the T cell clones shared between the liver and tumour. To this end, we performed single cell-resolved *in situ* spatial transcriptomics on liver and matched tumour tissue from three ChILI patients (P13, P26 and P31), using the Xenium platform. We designed padlock probes to target the CDR3 mRNA sequences of ten shared T cell clones: two from P13, four from P26 and four from P31 (**Supplementary Table 6**). We targeted hyperexpanded clones in the liver, defined as those with a frequency >1% of the total repertoire, as their increased abundance suggests antigen-driven selection and increases the likelihood of reliable detection and robust downstream analysis. These custom-designed CDR3 probes were added to a pre-designed probe panel (Human Immuno-Oncology Panel), along with probes for selected lineage-specific markers (**Supplementary Table 7**). From the liver, the entire biopsy was analysed. From the tumour, the analysed area was selected based on the previous BaseScope RISH to ensure the presence of shared T cell clones and included tumour and peri-tumour tissue (**Fig. 4E, Extended Data Fig. 9B**). Quality control showed a similar median gene and transcript count for the liver (gene: 81, transcript: 280) and the tumour (gene: 79, transcript: 184) samples (**Extended Data Fig. 10A-B),** and we observed no batch effects between sample types (**Extended Data Fig. 10C-D)**.

In the liver, 101,983 cells were segmented, and Louvain clustering identified 12 distinct cell phenotypes (**Fig. 5A**). These included two epithelial cell populations (hepatocytes and cholangiocytes), three stromal cell populations (stromal cells, liver sinusoidal endothelial cells [LSEC]-1 and -2) and seven immune cell populations (T cells, NK cells, B cells, plasma cells, macrophages, monocytes and pDCs). Cell annotation of the spatial transcriptomics data was confirmed by visually verifying the expected tissue morphology and by sequential mIF staining on the same tissue (**Extended Data Fig. 10E** and **Supplementary Table 8**). We observed consistent cellular composition in the Xenium and mIF analyses. Xenium data also showed a dominant infiltration of T cells, followed by macrophages, in ChILI liver (**Extended Data Fig. 10F**). In the three tumour samples, a total of 411,022 cells were segmented, and Louvain clustering assigned these cells to 11 main cell phenotypes (**Fig. 5A**). As in the liver samples, we identified three stromal cell populations (stromal cells, endothelial cells and desmin+ [DES+] cells) and six immune cells (T/NK cells, B cells, plasma cells, macrophages, neutrophils/MDSCs and pDCs). Additionally, we identified melanoma cells in the P13 tumour and breast cancer cells in the P31 tumour. The metastatic lung adenocarcinoma sample (P26) did not yield a distinct tumour cell cluster, likely due to low tumour cell infiltration in the tissue. Visual confirmation of cell distribution and phenotyping revealed the expected tissue morphology, which was again validated at the proteomic level by mIF staining with PCF (**Extended Data Fig. 10G-H**).

**Fig. 5.**
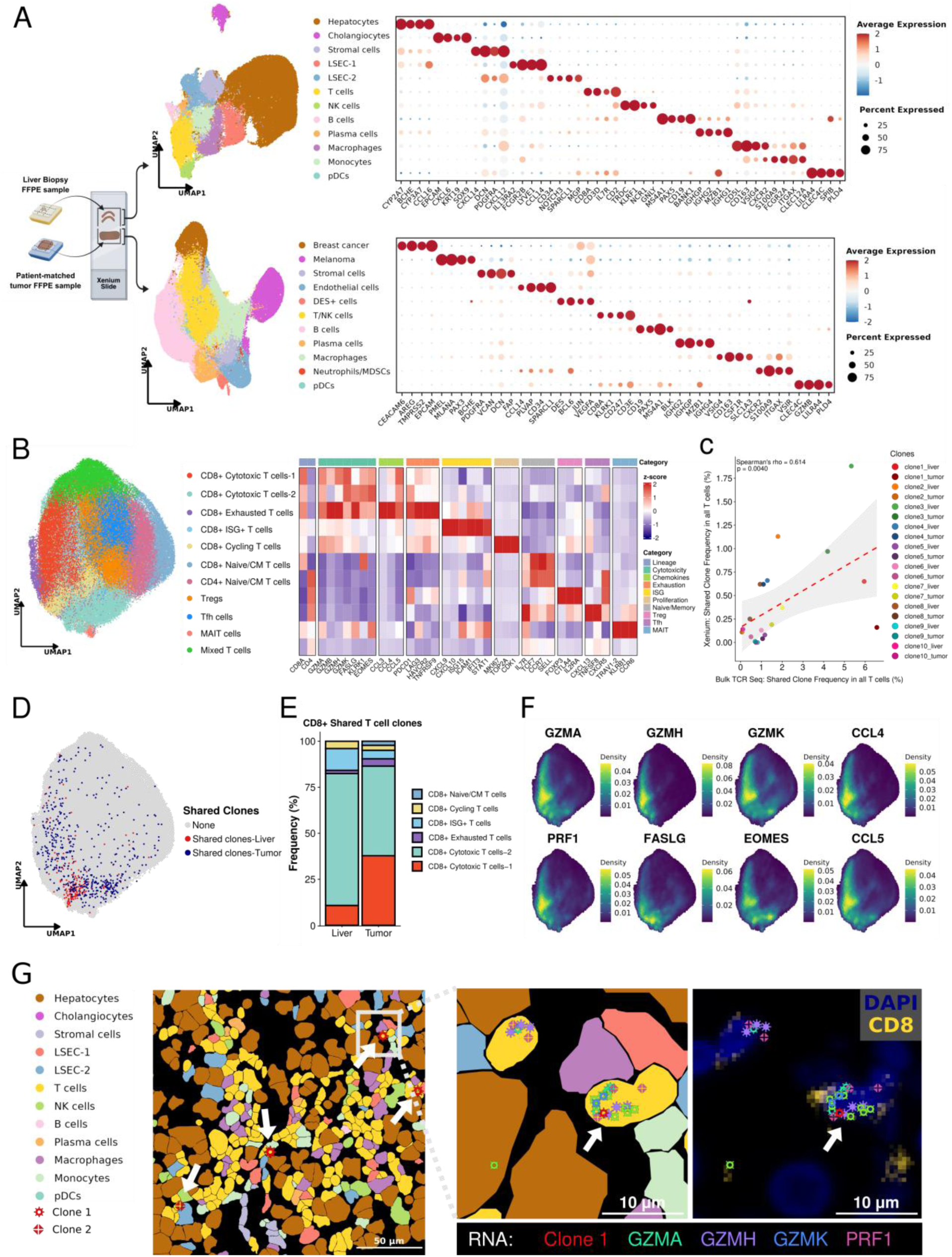
Shared T cell clones show cytotoxic phenotype in ChILI liver samples and patient-matched tumour samples. (A) UMAP projection of the main identified cell types (left) and dot plot showing the top 4 differentially expressed genes from each cell type (right), from spatial transcriptomics data of liver (top) and tumour (bottom) samples. (B) Left: UMAP projection of T cell subclusters combined from liver and tumour samples. Right: Heatmap shows the expression of selected genes in each T cell cluster. (C) Correlation plot of shared T cell clone frequency as determined by spatial transcriptomics and TCR-sequencing. Statistical test: Spearman’s correlation. (D) UMAP projection of shared T cell clones probed by our panel. Red dots represent shared T cell clones in the liver. Blue dots represent shared T cell clones in the tumour. Grey dots represent T cells not expressing a shared clone. (E) Phenotypic distribution of CD8+ shared T cell clones in the liver and tumour samples. (F) Visualization of cytotoxicity markers (*GZMA*, *GZMH*, *GZMK*, *PRF1*, *FASLG*, *EOMES*) and inflammatory chemokines (*CCL4* and *CCL5*) by Nebulosa density plots on the UMAP in D. (G) Exemplary visual representation of broad cell types and shared T cell clones within the liver of P13, as imaged with high-resolution spatial transcriptomics. Left: A region in the hepatic lobule infiltrated with T cells. White arrows indicate clone 1 and 2. White rectangle marks a region with clone 1. Scale bar = 50 µm. Centre: Magnified region with a clone 1 (white arrow), which expresses the cytotoxicity markers *GZMA*, *GZMH*, *GZMK* and *PRF1*. Right: Sequential mIF data on the same section showing CD8 expression on clone 1 (white arrow). Scale bar = 10 µm.

To enable a direct phenotypic comparison of T cells between liver and tumour, we merged the data corresponding to T cells from both tissues. We then subclustered the merged T cells and identified six CD8+ T cell populations: CD8+ cytotoxic T cells-1, CD8+ cytotoxic T cells-2, CD8+ exhausted T cells, CD8+ interferon stimulated genes (ISG)+ T cells, CD8+ cycling T cells and CD8+ naive/CM T cells; three CD4⁺ T cell populations: CD4+ naive/CM T cells, CD4+ follicular helper T cells [Tfh], and Treg, as well as MAIT cells and a mixed T cell population (**Fig. 5B**). To identify the shared clones, we examined the data from our clone-specific probes to determine which T cells from each patient expressed the relevant CDR3 transcripts. Any T cell with ≥1 such transcript was defined as belonging to a shared T cell clone. In the tumour, a few cells harboured transcripts from two distinct T cell clones and were excluded from the analysis (**Extended Data Fig. 11A**). In total, we found 187 T cells from shared clones in the liver and 522 in the tumour (33 and 155 for P13, 130 and 79 for P26, and 24 and 288 for P31, respectively). The frequencies of these shared T cells were between 0 and 1.88% in the liver and between 0.01 and 0.97% in the tumour, relative to the entire measured T cell compartment. The clone frequencies identified using spatial transcriptomics were generally lower than those obtained from bulk TCR sequencing (0.74–5.95% in the liver and 0.07–6.56% in the tumour). Nevertheless, we observed a moderate positive correlation between the frequencies of individual clones determined by the two methods (ρ = 0.614, p = 0.0040) (**Fig. 5C**), which supports the concordance of the two methods despite the differences in tissue section level and the distinct methodological principles underlying each approach. Additionally, we confirmed that the shared T cell clones were CD3 positive by performing immunofluorescence on the same slides used for spatial transcriptomics (**Extended Data Fig. 11B and Supplementary Table 8**).

Next, we analysed the phenotypes of the shared T cell clones. In keeping with our finding that CD8+ T cells are the most relevant T cell type in ChILI, 82.4% of the shared clones were classified as CD8+ T cells (**Fig. 5D**). The remaining shared clones were primarily various CD4+ T cell populations, but not MAIT cells. Most shared CD8+ T cell clones belonged to the CD8+ cytotoxic T cell population 1 or 2 and were found at similar frequency within all CD8+ T cells in the liver and the tumour (77.8% versus 81.1%) (**Fig. 5E**). Only a few of the shared CD8+ T cells wereCD8+ ISG+ T cells, and almost none were CD8+ exhausted T cells or CD8+ cycling T cells. The shared CD8+ cytotoxic T cells were characterised by strong expression of cytotoxicity markers *GZMA*, *GZMH*, *GZMK*, *PRF1, EOMES and FASLG* and pro-inflammatory chemokines *CCL4* and *CCL5* (**Fig. 5F**). Visual examination of liver and tumour samples confirmed the expression of these cytotoxicity-related genes, and we validated CD8 positivity at the protein level using mIF (**Fig. 5G, Extended Data Fig. 11C**). We further confirmed the cytotoxic T cell phenotype by sequentially applying mIF and BaseScope RISH staining on the same slide. Indeed, most shared CD8+ T cell clones showed GZMB expression in the liver of P13 and P26 (75% and 80%) (**Extended Data Fig. 12A-C, Supplementary Table 9).**

In summary, our analysis showed that most shared T cell clones in ChILI patients exhibited a cytotoxic CD8+ T cell phenotype and were present in the liver and the tumour at similar frequency.

### Shared T cell clones communicate with macrophages across the CCL5–CCR1 axis

To elucidate in more detail how shared T cell clones exert their cytotoxic effect in ChILI, we analysed the location of these shared clones and their spatial interactions with other cells of the liver. Since our focus in this analysis was on T cells in the liver microenvironment, we analysed only the three liver biopsies (P13, P26 and P31) and excluded the tumour samples. Subclustering of the spatial transcriptomics data from these samples yielded eight T cell populations, including four CD8+ T cell populations, one naïve/CM CD4+ T cell population, one Treg population, one MAIT cell population and one mixed T cell population (**Fig. 6A**). The CD8+ populations included: CD8+ cytotoxic T cells (which included some T cells with tissue residency features), CD8+ISG+ T cells, CD8+ T_RM_ cells and CD8+ cycling T cells (**Fig. 6A**). As before, 81.3% of the shared clones were CD8+ T cells (**Fig. 6B**), and predominantly CD8+ cytotoxic T cells, followed by CD8+ ISG+ T cells and a few CD8+ cycling cells (**Fig. 6C**). No shared T cell clone mapped to the CD8+ T_RM_ cell phenotype. Importantly, shared CD8+ T cell clones were more frequently of a cytotoxic T cell phenotype than the remaining CD8+ T cells (77.6% versus 44.2%; **Fig. 6C**) and showed strong expression of markers associated with cytotoxicity (**Fig. 6D)**.

**Fig. 6.**
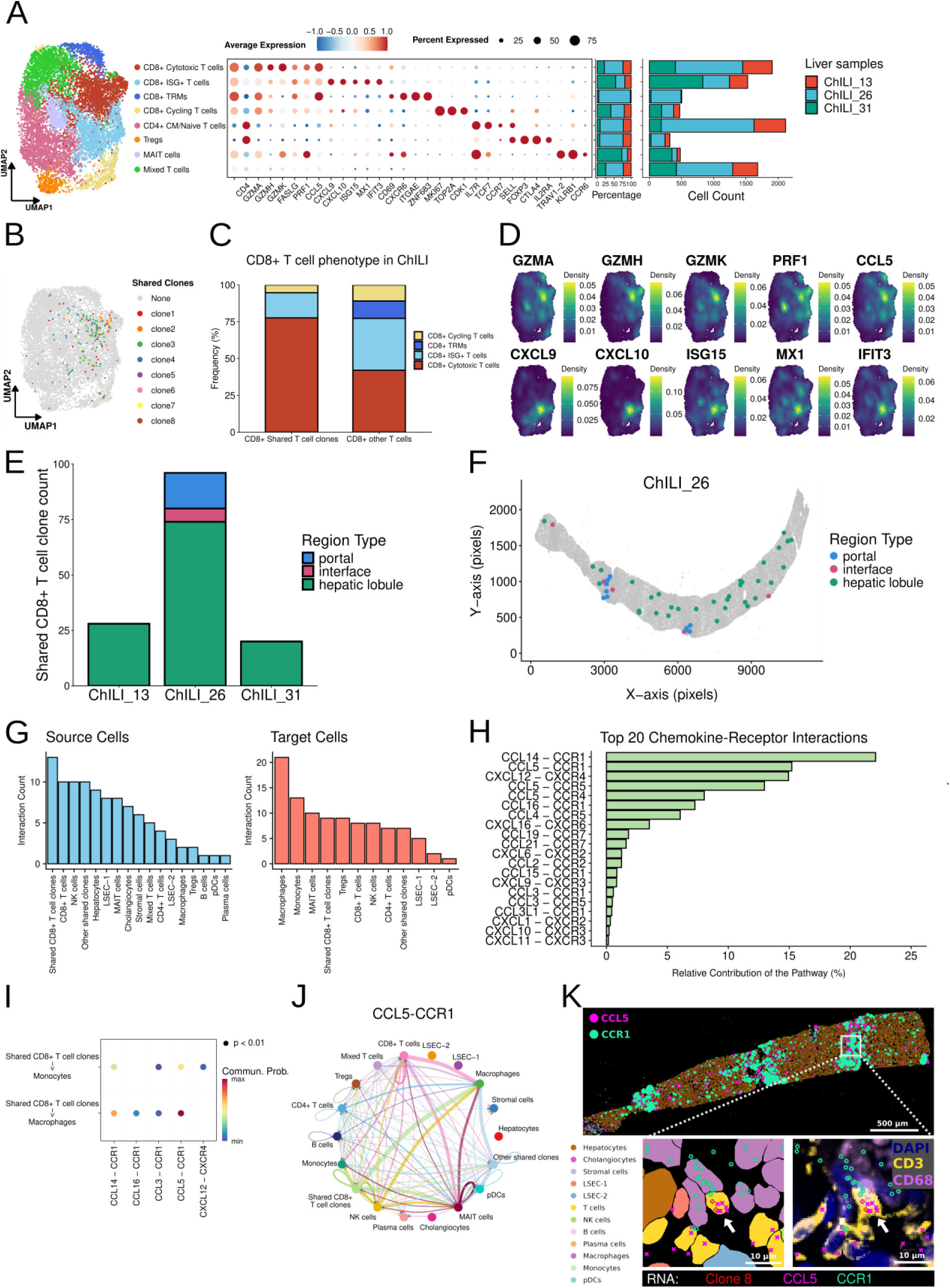
Shared T cell clones are distributed across different compartments of the liver and interact with other immune cells. (A) Left: UMAP projection of spatial transcriptomics data showing main T cell populations identified in liver samples from P13, P26 and P31. Right: Dot plot showing the selected genes defining each T cell population; bar plots show percentage and absolute counts of these populations in each patient. (B) Left: UMAP projection of shared T cell clones in the liver. Each colour represents a different clone. Grey colour shows T cells not expressing a CD3 clone in our panel. (C) Phenotypic distribution of CD8+ shared T cell clones in the liver. (D) Visualization of cytotoxicity markers (*GZMA*, *GZMH*, *GZMK*, *PRF1*, *CCL5*) and ISG markers (*CXCL9*, *CXCL10*, *ISG15*, *MX1*, *IFIT3*) by Nebulosa density plots on the UMAP in A. (E) Distribution of shared T cell clones in different liver compartments in each patient sample. (F) Representative dot plot from P26 showing shared T cell clone distribution in the liver. (G) Bar plots showing the source (left) and target (right) cell types corresponding to the top 100 strongest ligand–receptor pairs in a probabilistic interaction model of the spatial transcriptomics data. (H) Bar plots showing the relative contribution of the top 20 chemokine-receptor pairs contributing to the L-R network. (I) Bubble plot of significant ligand–receptor pairs between shared CD8+ T cell clones and monocytes and macrophages. Dot size encodes p-value and colour encodes communication probability (Commun. Prob). (J) Plot of the cell-cell communication network of the CCL5-CCR1 pathway. Nodes represent different cell types; lines represent interactions between two cell types. Line width encodes interaction strength; arrows indicate direction (source → target). (K) Exemplary visual representation of a liver region in P31 rich in CCL5-CCR1 expression. Top: Dots represent transcripts of CCL5 (magenta) and CCR1 (green). Rectangle marks the area magnified below. Bottom: Magnified area showing clone 8 and mapping of the indicated transcripts to broad cell types annotated from spatial transcriptomics data (left) and to IF images of the indicated markers.

Next, we localised the shared CD8+ T cells within the liver using masks for different liver compartments. Interestingly, the location of the shared T cell clones corresponded to the inflammatory pattern observed histologically. P26 displayed shared T cells in the lobules, as well as in the interface and portal tract (**Fig. 6E–F**). In contrast, P13 and P31 displayed shared T cells only in the lobules (**Fig. 6E**).

To gain a deeper insight into the interactions of shared CD8+ T cell clones with other immune cells, we analysed the transcriptomic data to assess ligand–receptor–mediated communication networks. Using probabilistic interaction modelling^25^, we ranked all predicted cell-to-cell interactions based on their communication probability. The shared CD8+ T cell clones emerged as the dominant cell type involved in intercellular communication. They ranked as the top source and fourth-ranked target cell type **(Fig. 6G)**. This suggests that shared CD8+ T cell clones exert a paracrine effect on other cells and also receive signals from their surroundings. Macrophages and monocytes were the top-ranked target cell populations, indicating that they communicate with the shared CD8+ T cell clones as well as with other cell types **(Fig. 6G)**. Given that chemokines are key players in cell-to-cell communications, we investigated chemokine ligand–receptor interactions in detail. We found that pathways involving CCR1 or CCL5 formed the largest proportion of the communication network **(Fig. 6H).** CCL5 and its cognate receptors CCR1 and CCR5 were expressed by T cell populations including shared CD8+ T cell clones and NK cells. In addition, CCR1 was expressed by macrophages, monocytes and pDCs, while CCR5 expression in non-T cells was restricted to pDCs. **(Extended Data Fig. 13A)**. Interestingly, the strongest ligand-receptor pair between shared CD8+ T cell clones and both monocytes and macrophages was CCL5-CCR1 **(Fig. 6I)**. Although we observed CCL2-expressing cells and CCR2+ monocytes **(Extended Data Fig. 13B)** as previous studies suggested ^9,11^, it did not form a significant T cell-monocyte/macrophage interaction. Shared CD8+ T cell clones were the primary source cells in the CCL5-CCR1 pathway, alongside CD8+ T cells and NK cells. In contrast, macrophages were the main target cells (**Fig. 6J**). Finally, we localized the expression of CCL5 and CCR1 in tissue. As expected, these transcripts co-localized with T-cell-rich immune infiltrates, where shared CD8+ cytotoxic T cell clones expressing CCL5 were in close contact with CCR1+ macrophages (**Fig. 6K**). These CCR1+ macrophages also expressed the T-cell-attracting chemokines CXCL9 and CXCL10, as well as the costimulatory molecules CD80 and CD86, indicative of an activated phenotype (**Extended Data Fig. 13C**). Notably, CXCL9 and CXCL10 transcripts were spatially co-localized with CCL5-and CCR1-enriched tissue regions, supporting coordinated chemokine-mediated crosstalk within these immune niches (**Extended Data Fig. 13D**).

These findings suggest that shared T cell clones interacting with macrophages and monocytes via the CCL5-CCR1 axis play a dominant role in ChILI.

## DISCUSSION

ICIs have changed cancer care across multiple tumour types. However, their benefit is counteracted by so-called irAEs, which impact therapeutic management.^1–3,5^ The liver is one of the organs most frequently affected by ICI therapy, yet the biological mechanisms underlying ChILI remain poorly understood.^26,27^

Here, we present the first single-cell, multiplexed proteomics atlas of ChILI in human liver biopsies. We demonstrated an expansion of CD8+ cytotoxic T cells alongside both CD68+CD163^Hi^ and CD68+CD163^Lo^ macrophages. Our analysis revealed that the interaction between CD8+ cytotoxic T cells and these two types of macrophages is a defining feature of ChILI. This indicates that CD8+ cytotoxic T cells and macrophages are key drivers of ChILI, as previously proposed in human studies^9^ and mouse models.^8–10^ Bulk transcriptomics further confirmed the multiplex proteomic results. It revealed robust immune activation and the enrichment of pathways involving key inflammatory mediators such as IFN-γ and TNF-α, as well as cytotoxicity.

Epitope sharing is thought to be one of the main mechanisms that trigger irAEs.^3,27,28^ In this way, neoantigens released from the tumour can lead to molecular mimicry, resulting in the activation of T cells that recognise self-antigens.^3,28^ Using bulk TCR sequencing of liver biopsies and matched tumour samples, we demonstrated for the first time that tumour–liver T cell clone sharing occurs in ChILI. Analogous observations have been made in irAEs of the lung, skin, and myocardium.^12,13,24^ A custom-made panel allowed us to track shared T cell clones specifically, using single-cell spatial transcriptomics. This novel approach has recently been used to track T cells with a specific TCR at a single-cell level to understand their function in more detail.^29–32^ We have now used this approach for the first time in irAEs caused by ICI. Interestingly, the phenotype of the shared CD8+ T cell clones was similar in the tumour and the liver. Most clones exhibited a cytotoxic phenotype. Importantly, the shared CD8+ T cell clones in the liver demonstrated more frequently a cytotoxic T cell phenotype than the remaining CD8+ T cells (77.3% versus 44.7%), suggesting that most shared clones are involved in liver injury. Single-cell spatial transcriptomics also enabled us to gain detailed insight into the immune microenvironment of ChILI. This indicated that CCL5-expressing shared CD8+ T cell clones interacted with CCR1+ macrophages. This suggests that the shared CD8+ T cell clones attract CCR1+ macrophages into proximity via the CCL5-CCR1 axis. In turn, these macrophages expressed the well-known T cell activators CXCL9 and CXCL10. Clonally expanded T cells, which expressed CCL5 and interacted with CCR1 or CCR5 have previously been described in patients with myocarditis undergoing ICI therapy.^33,34^ Similarly, single-cell RNA sequencing of synovial fluid from patients with ICI-induced arthritis revealed communication between exhausted CD8+ T cells and myeloid cells via the CCR1–CCL5/CCL3 and CXCL10–CXCR3 axes.^35^ Enhanced migration of peripheral monocytes towards CD8+ T cells was further attenuated through CCL5 blockade *in vitro*.^35^ Therefore, the CCL5-CCR1/CCR5 axis seems to be important also in other irAEs, besides ChILI. Previous studies on ChILI have highlighted the significance of the cellular communication between CD8+ cytotoxic T cells and CCR2+ monocytes and macrophages.^9,11^ We also identified CCR2+ monocytes within the microenvironment of shared T cell clones. However, these cells did not show a significant contribution to the communication network. This difference may be explained by variations between mice and humans and by our focus on shared T cell clones.

Our analysis also has implications for therapy. The main advantage of targeting a specific pathway, such as CCL5-CCR1, in irAEs, is the potential for tissue-specific immunosuppression. This could reduce organ-specific toxicity while preserving anti-tumour immunity. This contrasts with the current standard of care, which involves the use of non-selective agents, such as corticosteroids, that may compromise the efficacy of ICI in tackling tumours. As described above, the use of the CCR2–CCR5 dual inhibitor CVC to block the interaction between CD8+ T cells and macrophages has been demonstrated to reverse ICI-induced liver damage in a mouse model.^11^ Similarly, maraviroc, a well-established CCR5 inhibitor, which is already in clinical use for treating patients with HIV and which has relatively few known side effects, may be a pharmacological agent worth exploring for patients at risk of irAE with ICI.^36^ However, the question remains whether CCR5 or CCR1 blockade would also impair ICI anti-tumour efficacy, besides reducing irAEs. For instance, a recent study showed that the CCL5–CCR1/CCR5 axis promoted the polarization of macrophages towards an inflammatory M1 phenotype, which is critical for therapeutic efficacy.^37^

This study has limitations. First, its retrospective, cross-sectional design restricts conclusions to associations rather than causations. Second, although the shared T cell clones in the liver demonstrated a cytotoxic phenotype and underwent clonal expansion, indicating their involvement in ChILI, it remains unclear whether they initiated the disease or were involved propagating it. Third, as samples were collected around the time of toxicity, it was not possible to evaluate the longitudinal dynamics of clonal T cell populations. Prospective studies, involving serial sampling of peripheral blood at multiple time points alongside tissue-based analyses, are required to better understand the role of T cell clones in patients with ChILI.

In conclusion, our multimodal approach combining high-throughput spatial techniques revealed the phenotypic and spatial features of ChILI. Additionally, we analysed in detail hyperexpanded shared T cell clones present in both the livers and matched tumours of ChILI patients. Finally, we established an experimental and analytical workflow for tracking T cells *in situ*, that can be applied to a variety of biological questions.

## Data Availability

All data and code will be made available on Zenodo and Github upon publication.

## Abbreviations

ChILI: checkpoint inhibitor-induced liver injury
CN: cellular neighbourhood
FFPE: formalin-fixed paraffin-embedded
FOV: field of view
GZMB: granzyme B
ICI: immune checkpoint inhibitor
IF: immunofluorescence
IFN-γ: interferon-γ
IMC: imaging mass cytometry
irAEs: immune-related adverse events
LSEC: liver sinusoidal endothelial cell
mIF: multiplex immunofluorescence
PCF: phenocycler-fusion
pDC: plasmacytoid dendritic cells
RISH: RNA in situ hybridization
TCR: T cell receptor

## MATERIALS and METHODS

### Cohort design and ethical approval

In our cohort, most patients (80%) had grade 3 or 4 ChILI at biopsy. ChILI onset occurred at a median of 70 days after initiation of ICI therapy and a median of 26 days after the last ICI infusion. 43% (n=15) of the patients received immunosuppressive therapy at the time of biopsy - typically for only a few days - and all had elevated liver enzymes at the time of biopsy. All patients were negative by PCR or antibody testing for viral hepatitis (hepatitis A, B, C, D, and E), and none of the patients had a history of liver disease or alcohol abuse. Based on liver enzyme levels ^1^, 66% (n = 23) were classified as ChILI hepatitis, 20% (n = 7), as ChILI cholangitis, and 14% (n = 5) as mixed forms (**Supplementary Table 2**).

The study was approved by the Ethics Commission of Northern Switzerland (EKNZ; study ID: PB_2018-00308, 310/12). All tissue samples were collected as part of the routine diagnostic workup and selected retrospectively. The study was conducted according to the Declaration of Helsinki (1975).

### Clinical data collection

Diagnoses were established on clinical and morphological grounds according to current criteria. Comprehensive laboratory and serological data were collected that included liver function tests (ALT, AST, GGT, albumin, bilirubin) and autoantibodies including antinuclear antibodies (ANA), anti-smooth muscle antibodies (SMA), anti-mitochondrial antibodies (AMA), anti-actin antibodies (AAA), anti-liver/kidney microsome type 1 (LKM-1), anti-liver cytosol type 1 (LC-1), anti-soluble liver antigen/liver pancreas antigen (anti-SLA/LP), anti-neutrophil cytoplasmic antibodies (ANCA), serum immunoglobulin G (IgG) and ceruloplasmin levels (**Supplementary Table 2**). Autoimmune liver serology was evaluated according to local laboratory standards. Serological or PCR tests were performed for hepatitis virus A, B, C, D and E, Cytomegalovirus and Epstein–Barr virus.

### Morphological evaluation of patient samples

ChILI patient selection was based on a characteristic morphological spectrum previously described in the context of immune-related liver injury. Inclusion criteria required evidence of portal and lobular inflammation with hepatocyte damage, interface hepatitis, and the presence or absence of bile duct injury. Cases with clear histological evidence of alternative etiologies were excluded.

### Total RNA isolation from FFPE liver needle biopsy samples

FFPE tissue sections of 15-30 μm thickness were obtained from liver biopsy and tumour tissue blocks of ChILI patients and deparaffinized using xylene. Total RNA was extracted with AllPrep DNA/RNA FFPE Kit (Qiagen, 80234), according to the manufacturer’s protocol. The final RNA concentrations were quantified using the Qubit RNA HS Assay Kit (ThermoFisher, Q32852).

### Bulk RNA Sequencing preprocessing

Extracted FFPE RNAs from ChILI and normal liver samples were used for bulk RNA sequencing with the HTG Transcriptome Panel (HTG Molecular Diagnostics), as described before.^2^ The sequenced data was provided in FASTQ files format and aligned to the list of probes in the panel with the HTG EdgeSeq parser version 5.3.0.7148. The parsed data underwent post-sequencing quality control steps using the HTG Reveal software version 4.0.

### Differential gene expression analysis

Raw gene-level counts were analysed in R using edgeR (v4.2.0).^3^ Counts were imported into a *DGEList* object and lowly expressed genes were filtered prior to analysis. Library sizes were normalised using trimmed mean of M values (TMM), and dispersion parameters were estimated within edgeR. Differential expressions were tested using the quasi-likelihood framework (*glmQLFit* and *glmQLFTest*). Genes were called significant at FDR < 0.05 with an absolute log2 fold-change ≥ 0.5.

### Gene Set Enrichment Analysis

Gene set enrichment analysis (GSEA) was performed using the fgsea package in R (version 1.30.0). Genes were ordered by the log2 fold-change from differential expression analysis, and enrichment was tested using the MSigDB Hallmark collection.^4^ Significance was estimated from 10,000 permutations, and the results were corrected for multiple comparisons using the Benjamini-Hochberg method. Gene sets with a false discovery rate (FDR)-adjusted p-value < 0.05 were considered statistically significant.

### Deconvolution of immune cell type using MCP-counter

Deconvolution of immune cell subsets was computed using MCP-counter.^5^ Using TPM-normalised expression as input, we computed relative abundance for major immune populations - total T cells, CD8+ T cells, B cells, NK cells, macrophage/monocyte, myeloid dendritic cells, neutrophils and a cytotoxicity programme - as well as non-immune stromal compartments (endothelial cells, cancer-associated fibroblasts). Subsequent analyses concentrated on the immune cell population.

### Liver Biopsy Tissue Microarrays (TMA) for Phenocycler-Fusion (PCF) Staining

We reconstructed 8 TMAs by using the FFPE liver needle biopsy samples from ChILI and normal liver. We added one tonsil tissue in each TMA as a positive control for immune and epithelial markers. To construct TMA blocks, each tissue block was melted by using a tissue embedding station, and a maximum of 5 mm of tissue was cut from each biopsy sample as described before. ^2^

### Antibody panel for PCF

A 42-marker antibody panel was designed to target different non-immune liver cells such as hepatocytes (Arginase, Hep-Par-1, HNF4a), cholangiocytes (CK19, PanCK), endothelial cells (CD31), stromal cells (aSMA, Vimentin) and immune cell subtypes such as T-cells (CD45, CD3, CD4, CD8, CD45RO, FoxP3, CCR7, TCF-1, CD69, CXCR6), B-cells and plasma cells (CD20, CD38, CD138), granulocytes (CD66b, ECP, MPO), dendritic cells (CD11c, HLA-DR), macrophages (CD11b, CD68, CD163) and NK cells (NKG2A, CD57). In addition, cell state markers (PD1, PDL1, LAG3, Tim3, TOX, Granzyme B, IFN-γ, CD38, Ki67) were added to define the different states of immune cells. The markers with a low signal-to-noise ratio (TCRVa7.2, CCR2) were not included for further analysis, leaving 40 antibodies for final analysis. The Phenocycler-Fusion mIF experiment design, antibody clones and dilutions, and exposure times are shared in **Extended Data Fig. 1**, **Supplementary Table 3.**

### Multiplex Immunofluorescence (mIF) Staining with Phenocycler-Fusion System

5-µm thick sections were obtained from each TMA and two TMA sections were placed on the vendor indicated region on SuperFrost Plus Adhesion slides (Epredia, J1800AMNZ). As a result, four slides had two TMA sections. The sections were stained according to the standard protocol of the Phenocycler-Fusion system: The slides were deparaffinized in a dry oven at 60°C overnight and for 10 minutes in xylene, then rehydrated with a graded ethanol series (100%, 90%, 70%, 50%). For heat-induced epitope retrieval, slides were incubated in AR9 buffer (Akoya Biosciences, AR90001KT) for 20 minutes in a pressure cooker with high pressure. Slides were incubated in the hydration buffer for 4 minutes, in the staining buffer for 20 minutes, and then incubated with the primary antibody cocktail overnight at 4°C. The slides were fixed with 1.6% PFA for 10 minutes, 100% cold methanol for 5 minutes, and Phenocycler fixative reagent for 20 minutes. The stained slides were stored in the storage buffer at 4°C until the flow cell assembly. The flow cell assembly and image acquisition were performed the following day. The images were acquired with the Phenocycler-Fusion 2.0 Instrument in 16-bit format.

### Data pre-processing and cell segmentation

The preprocessed data was acquired in qptiff format from PhenoCycler-Fusion 2.0 software. The images were opened and inspected in QuPath version 5.1.0. ^6^ A pixel classifier was trained based on the DAPI signal with QuPath’s default multiscale features to annotate each liver biopsy individually, and biopsies were exported separately from QuPath in tiff format. The rolling ball background subtraction algorithm was performed for each channel with a rolling ball radius of 10.^7^ Segmentation was subsequently performed using the DeepCell/Mesmer algorithm implemented within the Steinbock pipeline.^8,9^ The following channels were used for segmentation: DAPI as nuclear marker and CD138, CD45, aSMA as cytoplasmic/membranous marker.

### Mask generation and integration

A hepatic lobule mask and a portal mask were created based on Arginase, CD138, HepPar-1, CD45, and aSMA stainings with default multiscale features of QuPath v5.1.0. ^6^ The hepatic lobule mask expanded 20 µm towards the portal compartment, and the portal mask expanded 20 µm towards the hepatic lobule, creating a 40 µm-wide interface zone between the hepatic lobule and the portal compartment. The resulting three annotation masks (hepatic lobule, interface zone, portal) were integrated into a SingleCellExperiment object ^10^ within the R environment.

### Broad phenotyping clustering

In order to identify broad phenotypes in our dataset (hepatocyte, cholangiocyte, stroma, vessel, B cell, CD4+ T cell, CD8+ T cell, eosinophil, myeloid/macrophage, neutrophil, NK cell and plasma cell), we performed unsupervised graph-based clustering (Jaccard index-based weights, Louvain algorithm for community detection). We used a modified R implementation of the PhenoGraph algorithm (Rphenoannoy; v.0.1.0) ^11^ (https://github.com/stuchly/Rphenoannoy). The following lineage specific markers were used: HepPar-1, Arginase, PanCK, CK19, Vimentin, aSMA, CD31, ECP, CD4, CD68, CD11b, CD138, MPO, CD8, CD3e, HLA-DR, CD11c, CD163, CD45, CD57, NKG2A, CD38, CD20, CD66b. Clustering with 3 different numbers of nearest neighbors (*k* = 90, 110, 130) were assessed. The selected number of neighbors was based on evaluation of the number of clusters, RMSD, and silhouette width. Subsequently, clusters were aggregated into metaclusters on the basis of marker and using the *clValid* (v.0.7) package in R using *k*-means clustering. This approach resulted in 12 phenotypes: hepatocyte, cholangiocyte, stroma, vessel, B cell, CD4+ T cell, CD8+ T cell, eosinophil, myeloid/macrophage, neutrophil, NK cell and plasma cell. Heatmaps visualizing mean marker expression values were generated with the R package *ComplexHeatmap*.

### T cell and macrophage/myeloid phenotyping clustering

To identify immune clusters of CD4+ T cell, CD8+ T cell, and myeloid/macrophage, we used a similar approach as outlined for broad cell phenotyping. For the CD4+ T cell compartment, we used the following state markers: TCF1, CCR7, CD57, FoxP3, TOX, CD45RO, LAG3, PD1, Ki67, Tim3, CD38, HLA-DR and defined a total of three CD4+ immune clusters: Treg, naïve-like TCF1+, and CD4+ T cell without specific marker profile referred to as CD4+ T cell. For the CD8+ T cell compartment, we used the following state markers: Granzyme B, TCF1, CCR7, CD57, TOX, CD45RO, LAG3, PD1, Ki67, Tim3, CD38 and defined a total of three CD8+ immune clusters: cytotoxic, central memory, and CD8+ T cell without specific marker profile referred to as CD8+ T cell. For the myeloid/macrophage compartment, three subtypes: CD68+ CD163^Hi^, CD68+ CD163^Lo^ and dendritic cell were defined using the following state markers: CD68, HLA-DR, CD11c, CD11b, PDL1, CD163. Each cluster was evaluated and assigned using heatmaps to visualize mean marker expression values of the specific state markers used for clustering. Clusters were aggregated into metaclusters on the basis of per-cluster marker means using the *clValid* (v.0.7) package in R using *k*-means clustering.

### Differential abundance analysis

Differential abundance testing was used to determine whether significant proportional differences exist for cell types between ChILI and normal liver tissues and between liver compartments. This testing is based on a negative binomial generalized linear model with quasi-likelihood functions as implemented in the EdgeR R package (version 4.3.0).^3^ Only cell types with a false discovery rate (FDR) < 0.1 were reported as significant after correcting the p-values for multiple testing with the Benjamin-Hochberg method. ^12^

### Cell-to-cell interaction

Pairwise cell-to-cell interaction evaluation for immune cell populations was performed using the *imcRtools* R/Bioconductor package (v.1.8.0) as described. For each image, we computed the average cell type-cell type interaction count (“Histocat” method) using a k-nearest-neighbour graph (k = 20) built from cell centroids. Then these interaction counts were compared against empirical null distribution based on cell type label permutation. Cell pair significance values were assigned as follows: 1 – interaction; 0 – no interaction; −1 – avoidance.

### Patch analysis

To evaluate spatial connectivity of cells, we used the *patchDetection* proposed by Hoch et al. ^13^ and implemented in the imcRtools package.^9^ A k-nearest-neighbour graph (k = 20) was built from cell centroids, and “patches” were defined as sets of mutually connected cells of the target phenotype (cytotoxic CD8⁺ T cells) on this graph. To ensure robustness, only components containing ≥5 cells were retained. The resulting patches provide a measure of spatial aggregation of cytotoxic CD8⁺ T cells.

### Cellular neighbourhood analysis

Cellular neighborhood (CN) analysis was conducted following the framework introduced by Schürch and colleagues^14^ and implemented in the imcRtools R/Bioconductor package.^9^ Briefly, we constructed a 20-nearest-neighbor graph using previously defined immune subtypes and broad phenotypes. For each cell, neighboring cells within a 60 µm radius were aggregated to define its neighborhood composition. CNs were then identified by clustering these neighborhood profiles; we selected the number of CNs (k) via a parameter sweep (k = 3–10) and chose the optimal k based on the average silhouette (k = 6). Finally, differential abundance testing (as described above) was used to assess whether specific CNs were enriched in ChILI compared with normal liver.

### T cell Receptor (TCR) Library Preparation and Next Generation Sequencing

FFPE RNAs extracted from liver biopsy and tumour tissues of ChILI patients were used to prepare the libraries for TCR sequencing. cDNA was synthesized from RNA by using the Ion Torrent NGS Reverse Transcription Kit (ThermoFisher, A45003). T-cell RNA was quantified by qPCR with TaqMan Gene Expression Assay, CD247 (20X, Hs00167901_m1). Final input was normalised to 1 ng of T cell derived RNA per sample. For samples with low T-cell RNA yield, maximum RNA input was loaded for library preparation. Next Generation Sequencing (NGS) libraries were prepared using the Oncomine TCR Beta-SR RNA Assay (ThermoFisher, A39359) according to manufacturer’s instructions. Amplified and barcode ligated libraries were purified with AMPure XP Reagent (Beckman Coulter, A63880) and quantified with the Ion Universal Library Quantitation Kit (ThermoFisher, A26217). The library pool was prepared by combining equal volumes of libraries at 50 pmol/L concentration and loaded into Ion 550™ Chip (ThermoFisher, A34537). The libraries were sequenced on an Ion GeneStudio S5 Prime Sequencer (ThermoFisher).

### TCR Sequencing Data Analysis

Read alignment to the International ImMunoGeneTics (IMGT) database and removal of low quality and off-target reads were performed using Ion Reporter Software (version 5.20) Oncomine TCR Beta-SR w1.4 RNA workflow (ThermoFisher). Identified T-cell clones with CDR3 amino acid sequence, variable and joining gene, productive read count number were obtained in csv files from the software (**Supplementary Table 4**). Shared T cell clones were defined as T cells with the same variable-joining gene and 100% CDR3 amino acid sequence match. The T cell clones were categorized into 5 groups according to their frequencies calculated by productive read counts: hyperexpanded (10^-2^ < x ≤ 1), large (10^-3^ < x ≤ 10^-2^), medium (10^-4^ < x ≤ 10^-3^), small (10^-5^ < x ≤ 10^-4^) and rare (10^-6^ < x ≤ 10^-5^). Similarity and diversity metrics were calculated as indicated below:

The *Jaccard Similarity Index (JSI)* is defined as:

JSI = |A ∩ B| / |A ∪ B|

Where:

– A is the set of unique TCR clones in liver biopsy sample.
– B is the set of unique TCR clones in matched tumour sample.
– |A ∩ B| is the number of shared TCR clones between the liver biopsy and matched tumour sample.
– |A ∪ B| is the total number of unique TCR clones across liver biopsy and matched tumour sample.

The *Clonality Score* is defined as:

Clonality = 1 - (H / log₂(N))

Where:

H is the Shannon entropy, calculated as:

*H = -Σ (pᵢ × log₂(pᵢ))*

pᵢ is the proportional frequency of the iᵗʰ unique TCR clone.

N is the total number of unique TCR clones in the repertoire.

### RNA In situ Hybridization and Dual RNA In situ Hybridization-Immunohistochemistry with BaseScope Assay

Two 1ZZ in situ hybridization probes targeting CDR3 sequence with flanking framework regions of the chosen hyperexpanded T cell clones of patient #13 and patient #26 were custom designed by Advanced Cell Diagnostics (patient #13 sequence no: 126244, patient #26 sequence no: 160175) (for probe sequences, **Supplementary Table 5**). The assay was performed according to the manufacturer’s standard protocol of RNA in situ hybridization (RISH) and dual RNA in situ hybridization/immunohistochemistry (dual RISH/IHC) using the BaseScope v2 RED Assay (ACD, Cat. No: 322910) and the RNA-Protein Co-detection Ancillary Kit (ACD, Cat. No: 323180). Main modifications for melanoma (Patient #13) and NSCLC (Patient #26) tissue samples include 15 minutes of target retrieval with co-detection target retrieval solution and 15 minutes of protease IV treatment. Anti-CD3 monoclonal antibody (1:4000, Cat. No: 17617-1-AP, ProteinTech) was used for the immunostaining and developed with an avidin/biotin-based peroxidase system (VECTASTAIN Elite ABC-HRP Kit, PK-6100). The experiment was performed on sequentially cut 4 µm sections of melanoma and NSCLC samples with the corresponding target probe, a positive control probe (PPIB) and negative control probe (DapB). The slides were scanned with Ventana DP200 scanner and the images were captured with Fiji extension of QuPath v5.1.0.

### Xenium In Situ

The standard tissue morphology quality control with H&E staining was previously performed before TCRβ-CDR3 sequencing; therefore, this step was not performed again before the Xenium in Situ experiment. Samples were processed at the Functional Genomics Center Zurich using Xenium v1 chemistry for FFPE samples with multimodal cell segmentation, following the manufacturer’s instructions. Briefly, samples underwent xylene-based deparaffinization before being transferred to Xenium cassettes. Decrosslinking was then performed, followed by an 18-hour probe hybridization step (for details on the gene panel and TCR clonotype probes, see the section “*Design of add-on custom gene panel and TCR clonotype probes*”). The following day, probe ligation and rolling-circle amplification were carried out. Subsequently, the samples were blocked and subjected to an 18-hour staining procedure with multimodal cell segmentation mix. This mix includes antibodies for staining cell membranes and cell interiors, and a universal interior label for ribosomal RNA. On the next day, autofluorescence quenching and nuclear staining were performed. Finally, the processed Xenium slides were analysed using the Xenium Analyzer. All experimental procedures strictly adhered to the protocols provided by 10x Genomics.

### Design of add-on custom gene panel and TCR clonotype probes for Xenium In Situ

The 10x Genomics pre-designed Xenium Human Immuno-Oncology Profiling Panel (380 genes, Cat. No: 1000654) was supplemented with additional genes (predesigned panel ID: VVWA4W, Cat. No: 1000651) chosen to characterise the ChILI liver and tumour samples and hyperexpanded T cell clones (for the complete gene list of add-on custom gene panel: **Supplementary Table 7**). Since the pre-design panel is missing the genes expressed in parenchymal cells of the liver, we added probes to identify transcripts highly expressed in hepatocytes (*HEPN1*, *GLUL*, *BCHE*, *CYP2A7*, *CYP3A7*) and cholangiocytes (*KRT7*, *KRT19*). The custom add-on panel also expanded the list of targeted transcripts which are expressed in immune and stromal cells such as T cells, NK cells, B-cells and plasma cells, fibroblasts, endothelial cell, monocyte and macrophages. TCR probes were designed by the 10X Genomics Applied Bioinformatics team for CDR3 sequences corresponding to each shared hyperexpanded T cell clone identified in the liver explant (Cat. No: 1000664, for the complete TCR probe list, see **Supplementary Table 6**). Consequently, probes were successfully generated for 10 distinct shared T cell clones, with one probe per clone.

### Xenium In Situ Data Analysis

Spatial transcriptomic data was generated using the 10x Genomics Xenium platform (instrument software version 3.2.1.2, analysis version xenium-3.2.0.7). Analysis was performed with R version 4.4.2 using Seurat v5 as the primary analytical framework. The images were obtained with Xenium Explorer version 3.2.0.

### Cell segmentation of Xenium In Situ

Cell segmentation was performed based on multimodal cell segmentation immunofluorescence stain with standard Xenium segmentation algorithm provided by 10X Genomics that targets nucleus (DAPI), membrane (ATP1A1, E-Cadherin, CD45, 18S Ribosomal RNA) and cell interior (18S Ribosomal RNA, alphaSMA, Vimentin).

### Cell type annotation of Xenium In Situ

For the liver dataset, cells with less than 10 transcripts were filtered out. Raw counts were log-normalised. Given the limited size of the Xenium gene panel (∼350 genes), all available genes were used for scaling and PCA rather than selecting a subset of highly variable features. A nearest neighbour graph was constructed on the top 20 principal components and Louvain clustering was performed with a resolution of 0.3, selected based on silhouette score analysis across a range of resolutions and cluster stability assessed via cluster tree analysis. Uniform Manifold Approximation and Projection (UMAP) was used to visualize the cells on two dimensions. Cell type identities were validated through the expression of canonical marker genes and marker expression in https://www.livercellatlas.org/ and in MacParland et al. ^15^

For the tumour dataset, cells with less than 15 transcripts were filtered out. Raw counts were log-normalised. As above, all available genes were used for scaling and PCA. A nearest neighbour graph was constructed on the top 20 principal components and Louvain clustering was performed with a resolution of 0.4, selected based on silhouette score analysis across a range of resolutions and cluster stability assessed via cluster tree analysis. Uniform Manifold Approximation and Projection (UMAP) was used to visualize the cells on two dimensions. Cell type identities were validated through the expression of canonical marker genes.

To characterize T cell subpopulations, cells annotated as “T cells” in the liver dataset and “T/NK cells” in the tumour dataset were extracted and merged into a unified object. Rather than relying on all available panel genes, a curated gene list of 225 genes enriched in T/NK cells relative to all other cell types was derived using FindMarkers() in each dataset independently, followed by removal of genes associated with contaminating populations including myeloid, B, dendritic, stromal, and tumour cells. PCA was performed using this curated gene list and the top 20 principal components were retained. A nearest neighbour graph was constructed and Louvain clustering was performed at a resolution of 0.5, selected based on silhouette score analysis across a range of resolutions and cluster stability assessed via cluster tree analysis. UMAP was used to visualize the cells on two dimensions. Cell type identities were validated through the expression of canonical marker genes.

CCR1 status was assigned to each cell based on raw expression values; macrophages with expression greater than zero were classified as CCR1+, and the remainder as CCR1−.

### T cell clonotype analysis with Xenium In Situ

Shared clones were classified as positive based on the detection of at least one transcript per clone. Cells exhibiting signals from more than one clone (doublets) were excluded from being annotated as shared clones. Following cell type annotation, clones identified as non-T cells were also excluded from the shared clone list. Finally, shared clones annotated as T cells were visually validated at the proteomic level using CD3 staining from Phenocycler-Fusion mIF on the same slide. This workflow is summarized in **Extended Data Fig. 11A**.

### Post-Xenium mIF staining with Phenocycler-Fusion System and data analysis

The Xenium stained slides were stained according to the standard protocol of the Phenocycler-Fusion system as described above in the section “*Multiplex Immunofluorescence (mIF) Staining with Phenocycler-Fusion System”* with slight modifications. For heat-induced epitope retrieval, slides were incubated in AR9 buffer (Akoya Biosciences, AR90001KT) for 15 minutes in a pressure cooker with low pressure. The antibody panel was designed to generate tissue masks and identify main cell lineages. The post-Xenium PCF antibody panel is shared in **Supplementary Table 8**.

The images from Xenium in situ and PCF were aligned with Warpy 0.3.1 in QuPath v5.0. The Xenium morphology_focus_0001.ome.tif was chosen as the base image and PCF image was upscaled x2.38 based on the image pixel size ratio between Xenium and PCF images. Images were co-registered with affine transformation (pixel size: 0.25). Then a tissue mask was created based on DAPI, Arginase and CD138 stainings of PCF with default multiscale features of pixel classifier in QuPath v5.0. Next, A hepatic lobule mask and a portal mask were created based on Arginase, CD138 and CD45 stainings with default multiscale features with pixel classification. The hepatic lobule mask expanded 20 µm towards the portal compartment, and the portal mask expanded 20 µm towards the hepatic lobule, creating a 40 µm-wide interface zone between the hepatic lobule and the portal compartment. The PCF mIF images and three annotation masks (hepatic lobule, interface zone, portal) were exported with these specifications to make it compatible to Xenium Explorer software: compression type: ZLIB (lossless), pyramidal scale: 2.0, tile size: 1024 px and parallelize export. The resulting ome.tiff files were uploaded to Xenium Explorer version 3.2.0 for visualization. The cells in the portal and interface zones were manually selected by visual confirmation and exported as cell stats csv file containing specific cell IDs. Region specific cell IDs were used to integrate the spatial information into the Seurat object within the R environment.

### Cell–cell communication analysis (CellChat) on Xenium data

CellChat v2^16^ was used to infer intercellular communication from Xenium spatial transcriptomic data. For each ligand–receptor (L–R) pair and sender–receiver metacluster pair, CellChat computes a communication probability that integrates ligand and receptor expression across the sender and receiver groups. We estimated overexpressed ligands, receptors, and L–R pairs using CellChat’s built-in models with default scaling, and computed edge weights as the product of communication probability and the fraction of sending/receiving cells expressing the relevant partners.

### Combined Multiplex Immunofluorescence (mIF) Staining with Phenocycler-Fusion System and RNA In situ Hybridization with BaseScope Assay

Tissue blocks of tumour samples were scored with a clean scalpel and a 5-µm thick tissue section was obtained from the whole liver biopsy and chosen ROI of tumour samples of patient #13 and patient #26. The liver biopsy and tumour section from each patient were placed in the same slide. The tissues were stained according to the standard protocol of the Phenocycler-Fusion system as described above in the section “*Multiplex Immunofluorescence (mIF) Staining with Phenocycler-Fusion System”*. The antibody panel is shared in **Supplementary Table 9**. Following mIF staining, the sample slide-flow cell assembly was separated after overnight incubation in xylene. Then, the standard protocol of BaseScope RISH assay was performed as described above in the section *“RNA In situ Hybridization and Dual RNA In situ Hybridization-Immunohistochemistry with BaseScope Assay”* without the target retrieval step. The stained slides were scanned with VENTANA DP 200 slide scanner with 40X objective (Roche).

### Data visualization

Cell type–annotated segmented images were generated using the cytomapper R package (version 1.2.0).^17^ Dimensionality reduction plots (UMAPs) were produced with dittoSeq (version 1.2.6).^18^ Additional visualizations were created using ggplot2 (version 3.3.7) and ComplexHeatmap (version 2.20.0).^19^ Schematic illustrations were designed with BioRender (https://biorender.com/).

### Statistical analysis

The normality of numerical clinical parameters was assessed with visual (histograms, probability graphs) and analytical methods (Shapiro-Wilk test, skewness–kurtosis). For normally distributed variables, an independent two-sided T-test was conducted, whereas non-normally distributed variables were analysed using the Mann-Whitney U test. Categorical variables were compared using Fisher’s exact test. Comparison analyses were conducted using the Wilcoxon signed-rank test. To account for multiple hypothesis testing, we applied the Benjamini-Hochberg method to correct P values, and the false discovery rates (FDR q-values). Statistical significance was determined using a p-value threshold of < 0.05.

---

**Extended Data Figure 1.**
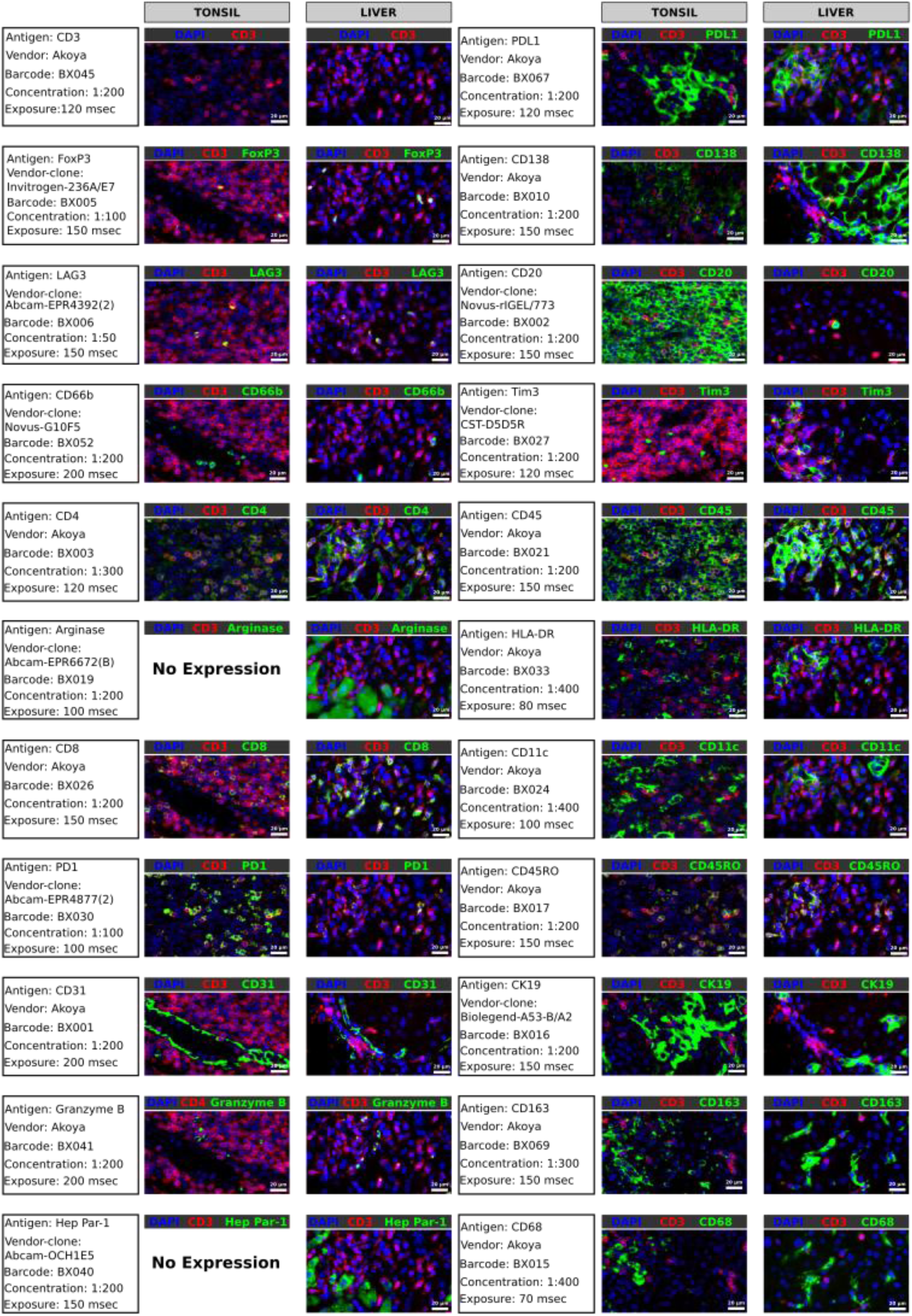
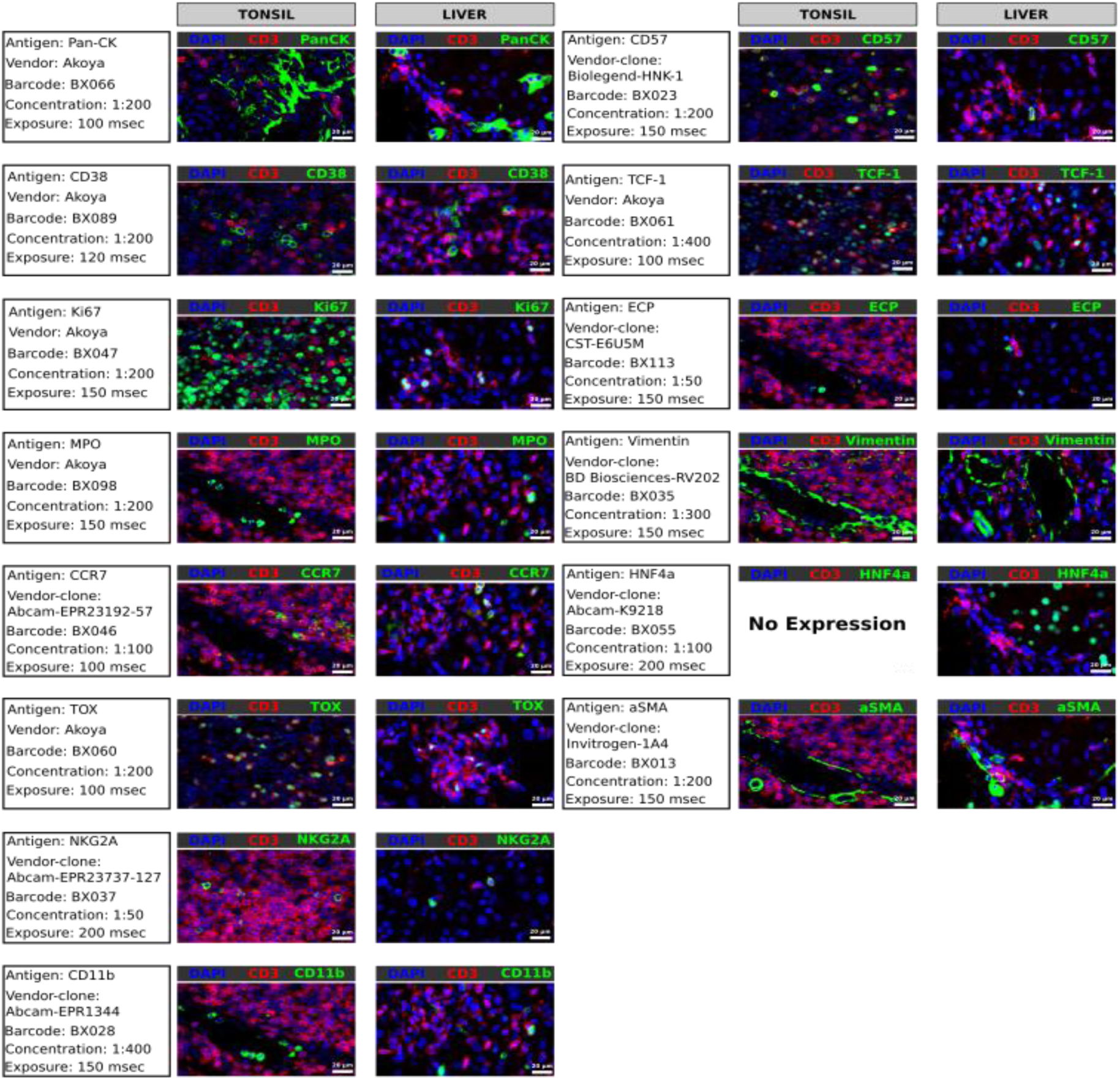
Antibody staining for mIF panel on human tonsil and liver tissues. Representative images demonstrating immunofluorescence staining of selected antigens included in the Phenocycler-Fusion mIF panel across tonsil (left column) and liver (right column) tissues. Antibodies were validated for specificity, signal intensity, and tissue compatibility. Each row shows staining for a given antigen (green), CD3 as the T cell lineage marker (red) and DAPI (blue) as a structural reference. Acquisition details including antibody clone, vendor, barcode, dilution, and exposure time are listed for each antigen. Scale bar = 20 μm.

**Extended Data Figure 2.**
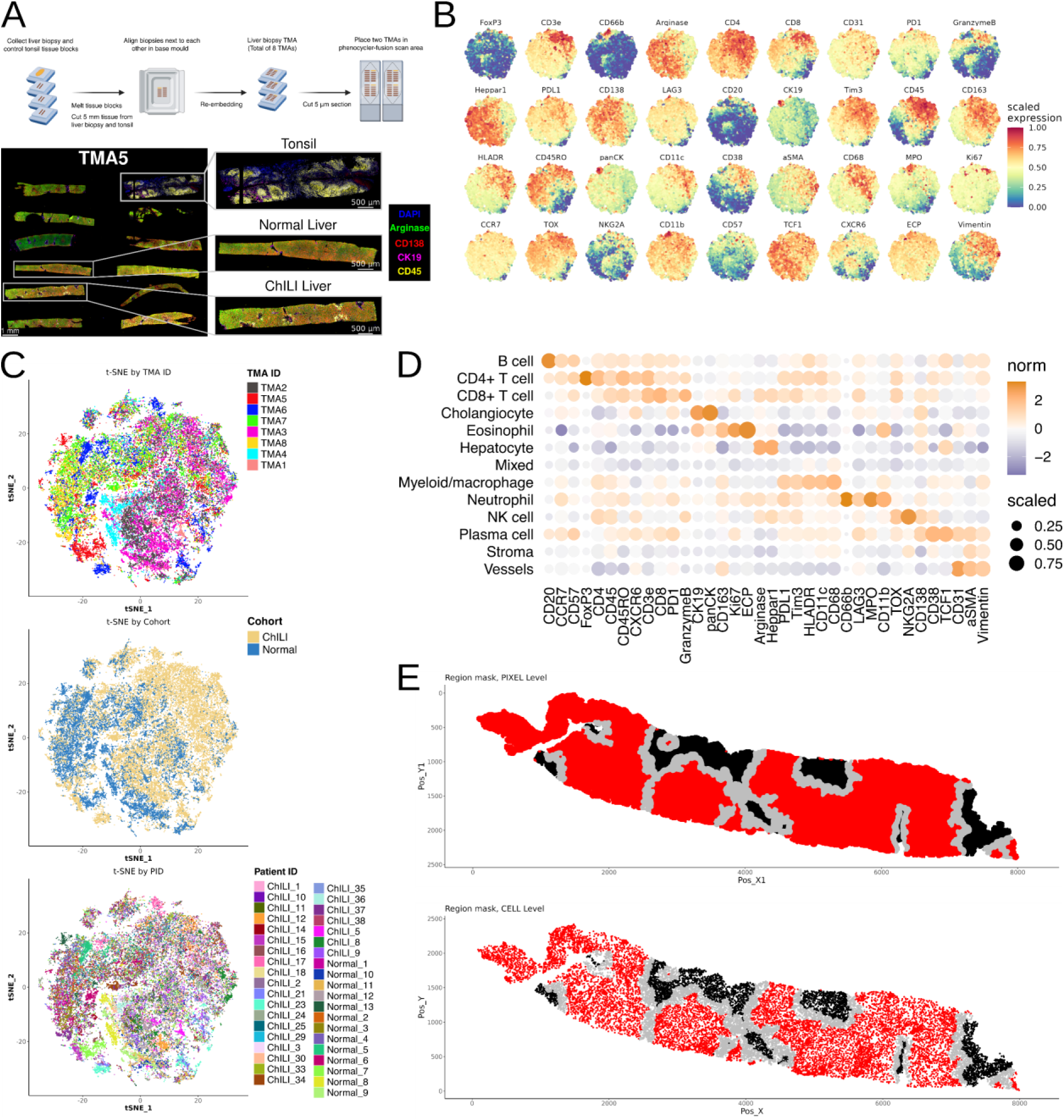
Single-cell, highly multiplex map of ChILI. (A) Top: schematic representation of the liver biopsy TMA preparation procedure. Bottom: whole-slide image of a liver biopsy TMA (left, scale bar= 1 mm) and a magnified view of a liver biopsy samples from normal and ChILI patient and tonsil sample (right, scale bar= 500 μm). (B) t-SNE representation of all single cells across each marker used in the mIF analysis; each plot shows the normalised expression of each marker. (C) t-SNE representation of all single cells colored by TMA (top), cohort (middle), and individual patient ID (bottom). (D) Dot plot showing mean scaled expression of all markers (x-axis) across annotated cell types (y-axis). Dot size indicates the fraction of cells expressing the marker, while colour intensity reflects scaled mean expression. (E) Pixel-level (upper panel) and cell-level (lower panel) dot plot of one representative liver biopsy (patient ChILI_25). Pixels and cells are labelled according to their region localization, portal (black), interface (grey), and lobular (red).

**Extended Data Figure 3.**
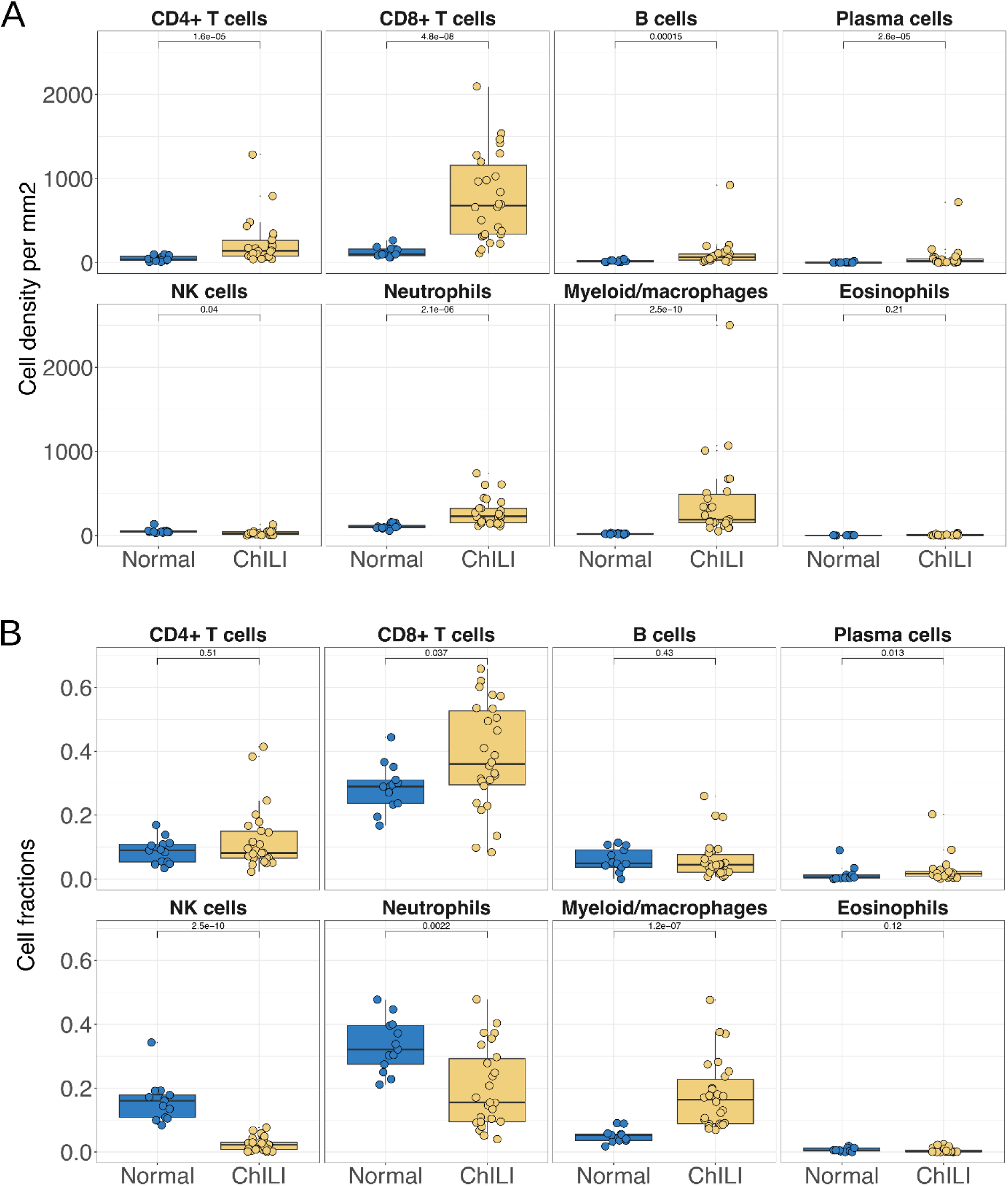
Abundance of broad immune phenotype across conditions. (A) Boxplots of absolute immune cell densities (cells/mm²) per subtype across cohorts (Normal vs ChILI). (B) Boxplots of absolute immune cell fraction per subtype across cohorts (Normal vs ChILI). Statistical comparisons were performed using Wilcoxon rank-sum test with Benjamini-Hochberg correction; adjusted p-values are indicated.

**Extended Data Figure 4.**
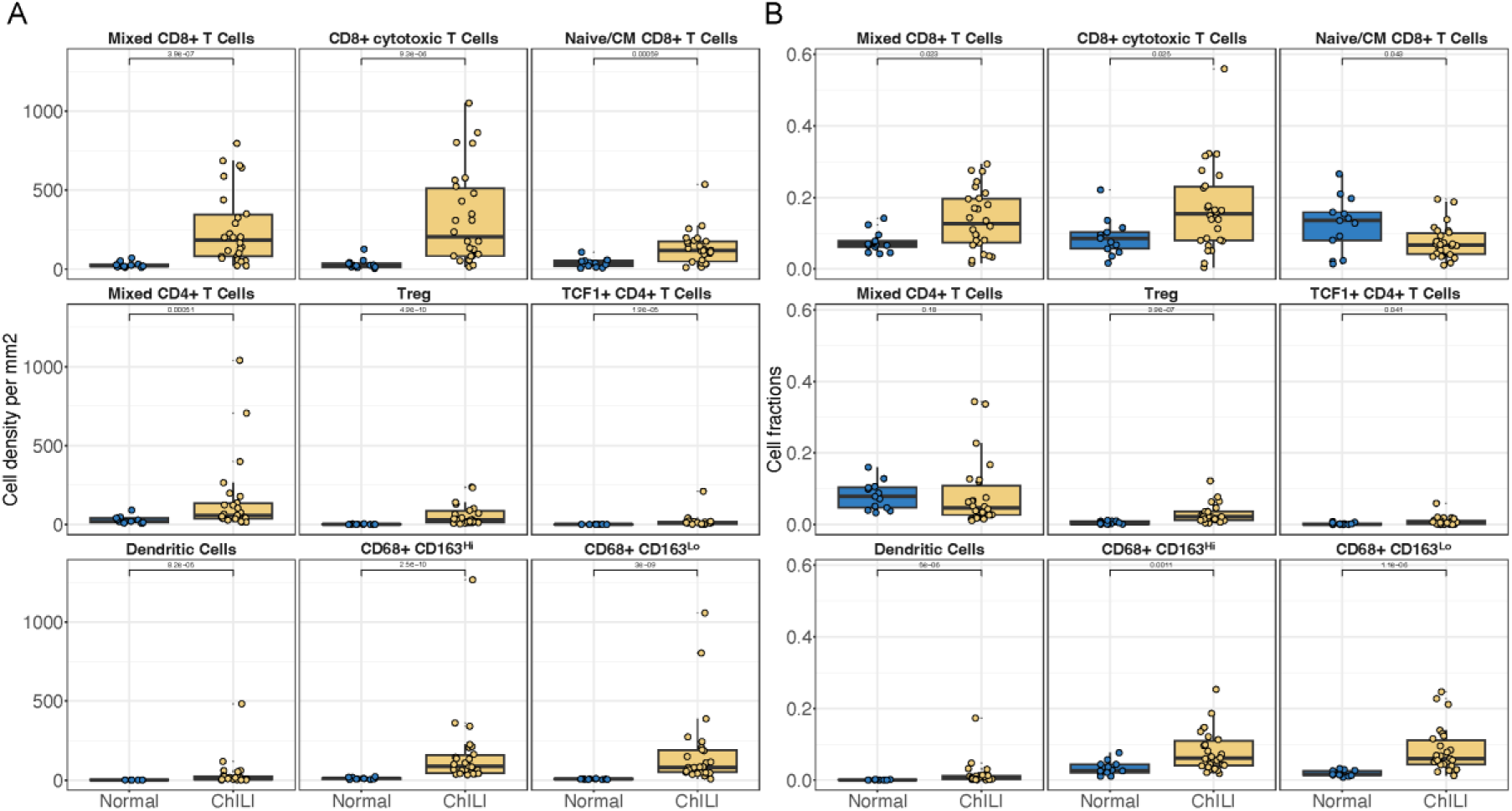
Abundance of immune clusters across conditions. (A) Boxplots of absolute immune cell densities (cells/mm²) per subtype across cohorts (Normal vs ChILI). (B) Boxplots of absolute immune cell fraction per subtype across cohorts (Normal vs ChILI). Statistical comparisons were performed using Wilcoxon rank-sum test with Benjamini-Hochberg correction; adjusted p-values are indicated.

**Extended Data Figure 5.**
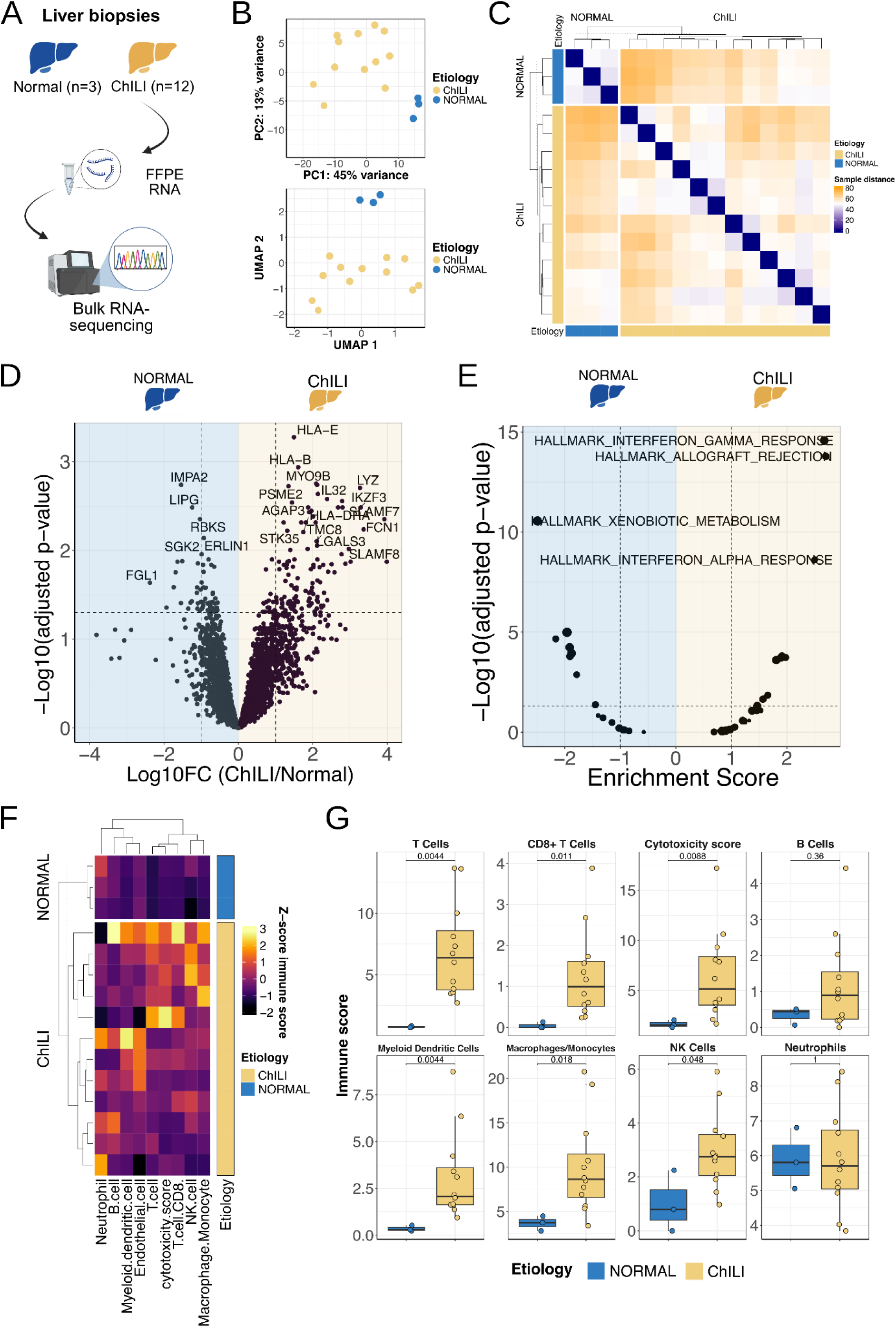
Differential gene expression from bulk transcriptomic in ChILI. (A) Overview of the experimental workflow showing bulk RNA-sequencing on FFPE liver biopsy specimens from patients with ChILI (n = 12), and normal liver controls (n = 3). (B) Principal component analysis (PCA, top) and Uniform Manifold Approximation and Projection (UMAP, bottom) based on transcriptomic profiles. Each dot represents one patient. (C) Heatmap of sample-to-sample transcriptomic similarity based on Spearman correlation distance from the 2’000 most variable transcripts. Sample clustering by etiology. (D) Volcano plot showing differentially expressed genes between normal liver and ChILI. Genes with adjusted p-value < 0.05 and absolute log₁₀ fold change > 1 are considered significant. Dashed lines: logFC and p-value thresholds. Each dot represents one transcript. Upregulated immune-related genes in ChILI are highlighted in bold. (E) Dot blot showing GSEA comparison between normal liver to ChILI using the MSigDB Hallmark gene sets. X-axis: enrichment score, Y-axis: -log₁₀ adjusted p-value, dashed lines: enrichment and p-value thresholds. (F) Heatmap of immune scores per sample derived from bulk RNA-seq using MCP-counter and a predefined cytotoxicity signature. Scores are Z-scaled by row. Columns correspond to individual liver biopsies (ChILI, n = 12; normal, n = 3) and are hierarchically clustered (Ward.D2; Euclidean distance). (G) Box-and-jitter plots show per-sample immune scores for eight cell types/signatures. Each dot is one sample; boxes indicate the median and interquartile range with whiskers extending to 1.5× IQR. Statistical test: two-sided Wilcoxon rank-sum test after multiple testing BH.

**Extended Data Figure 6.**
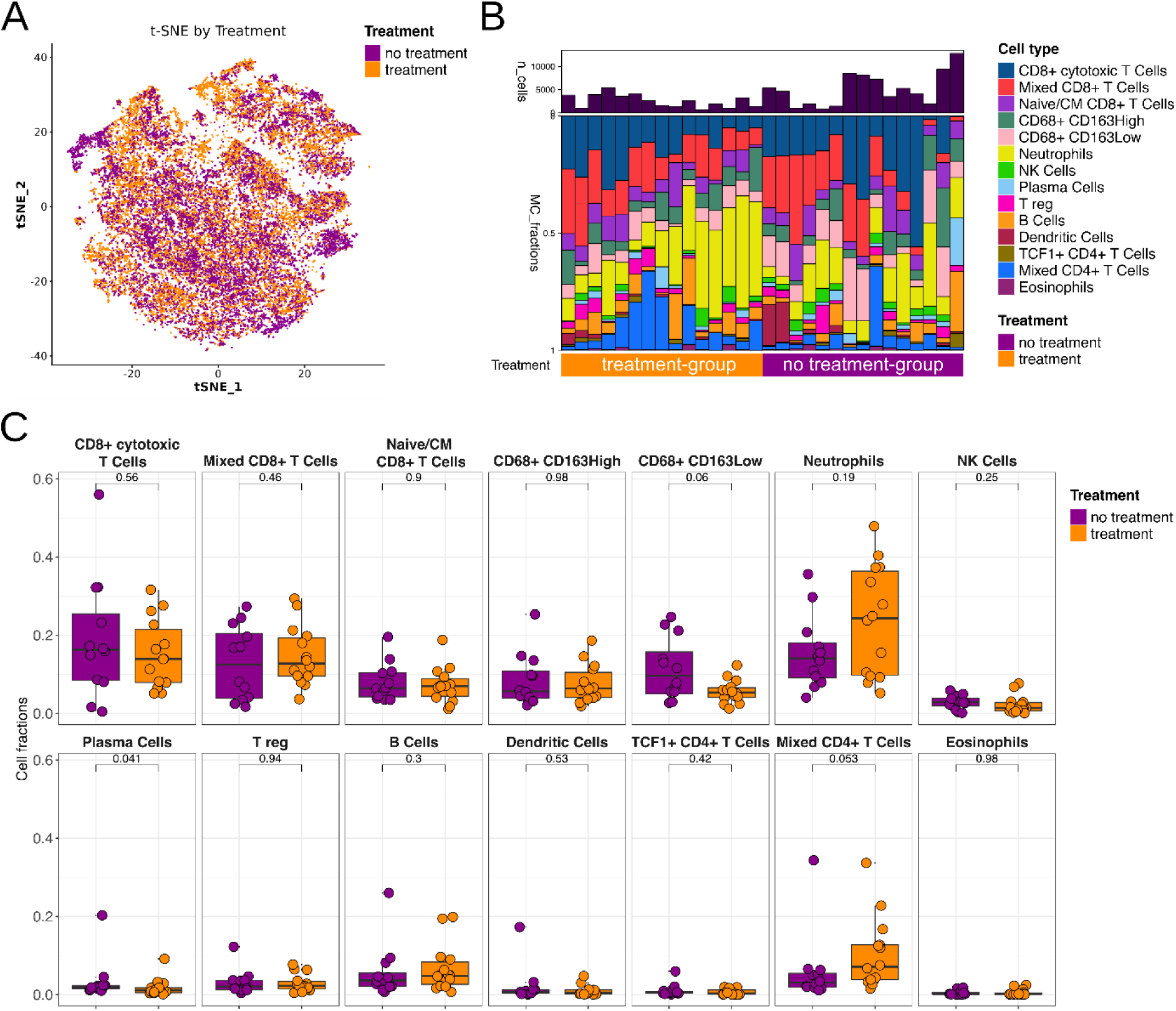
Immune composition of ChILI samples according to immunosuppressive treatment. **(A)** t-SNE embedding of all cells colored by treatment status showing substantial overlap between groups. **(B)** Stacked bar plots showing the relative abundance of the indicated immune cell populations in each sample; the upper bar plot indicates total cell numbers per sample, and the lower annotation denotes treatment status. **(C)** Boxplots with individual samples overlaid comparing cell-type fractions between treated and treatment-naïve samples. Each dot is one sample; boxes indicate the median and interquartile range with whiskers extending to 1.5× IQR. Statistical test: two-sided Wilcoxon rank-sum test after multiple testing BH.

**Extended Data Figure 7.**
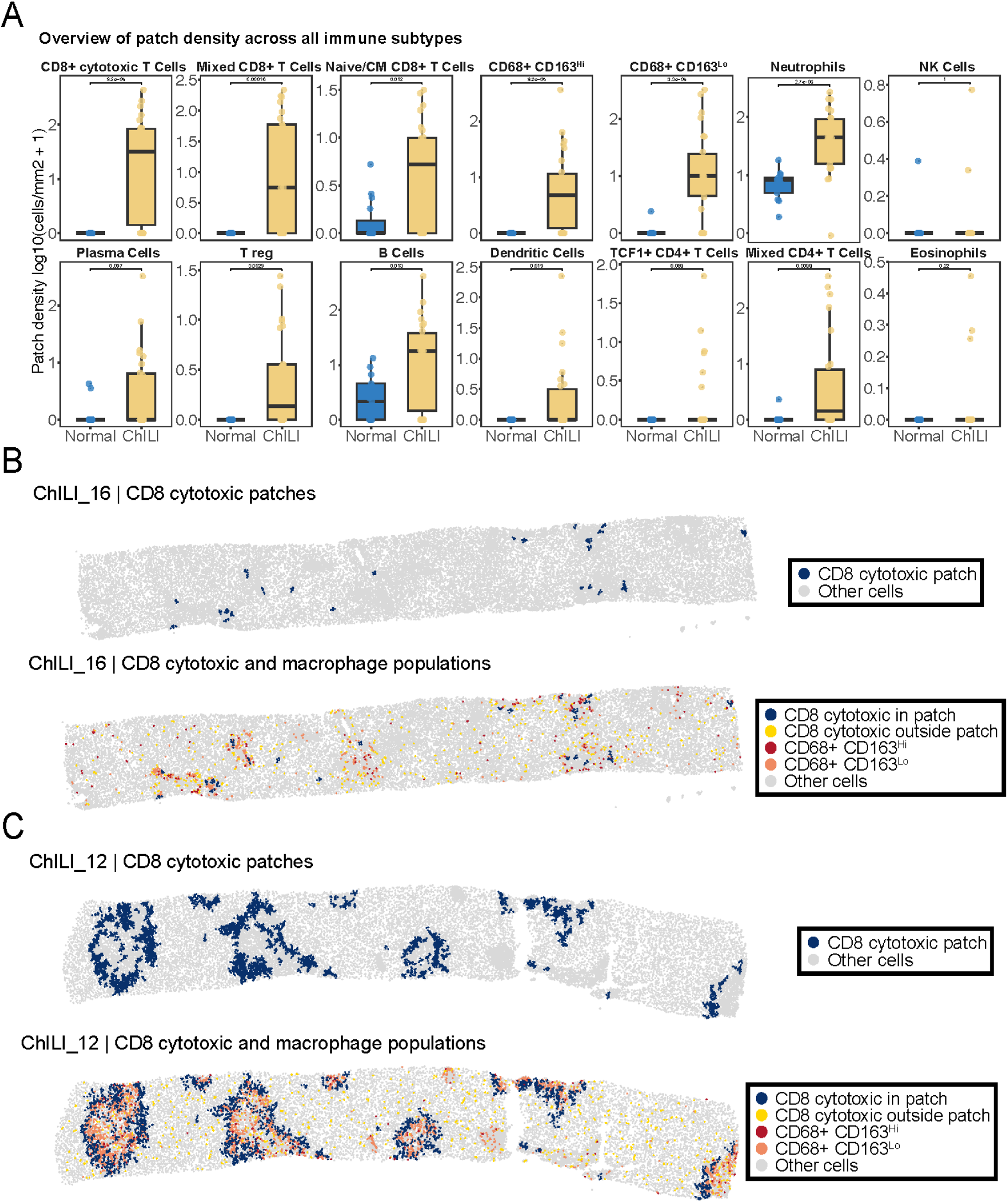
Patch density across immune subtypes in ChILI. (A) Patch density of the indicated immune subtypes in normal and ChILI liver biopsies, shown as log10 (cells/mm² + 1). Each dot represents one biopsy; boxes indicate the median and interquartile range; p values are shown above each panel. (B,C) Representative spatial maps from samples ChILI_16 (B) and ChILI_12 (C). Upper panels show CD8+ cytotoxic T cell patches (dark blue) across the full tissue section; all other cells are shown in grey. Lower panels show the same sections highlighting CD8+ cytotoxic T cells within patches (dark blue), CD8+ cytotoxic T cells outside patches (yellow), CD68+CD163^Hi^ macrophages (red), CD68+CD163^Lo^ macrophages (orange), and all other cells (grey).

**Extended Data Figure 8.**
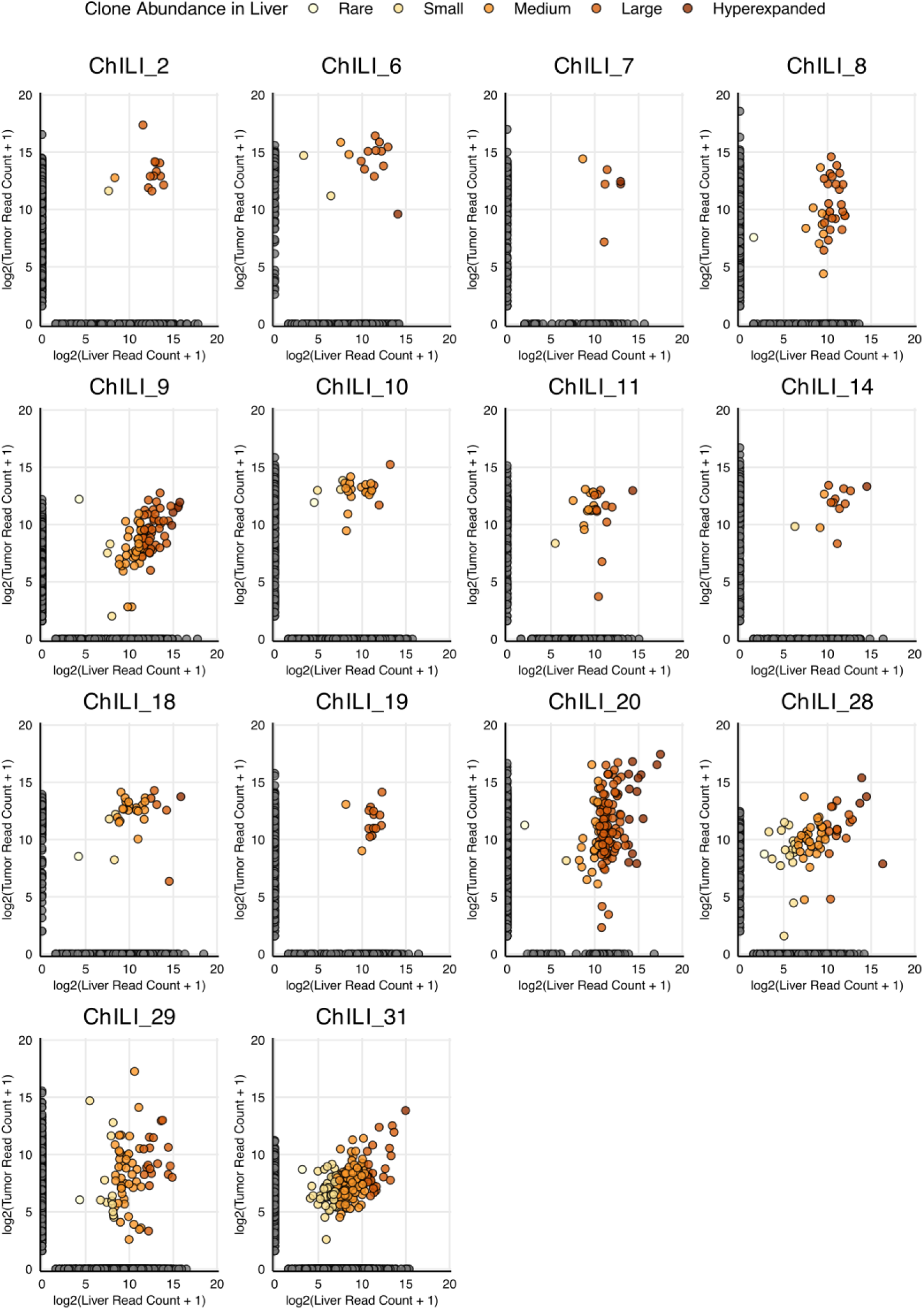
Clonal abundance of T cell clones shared in paired liver-tumour biopsies from each ChILI patient. Log2 normalised productive read count of T cell clones from all patients included in the study. Each dot indicates a clone, and shared clones were coloured based on clonal abundance. Gray colour indicates the remaining clones unique to either liver or tumour.

**Extended Data Figure 9.**
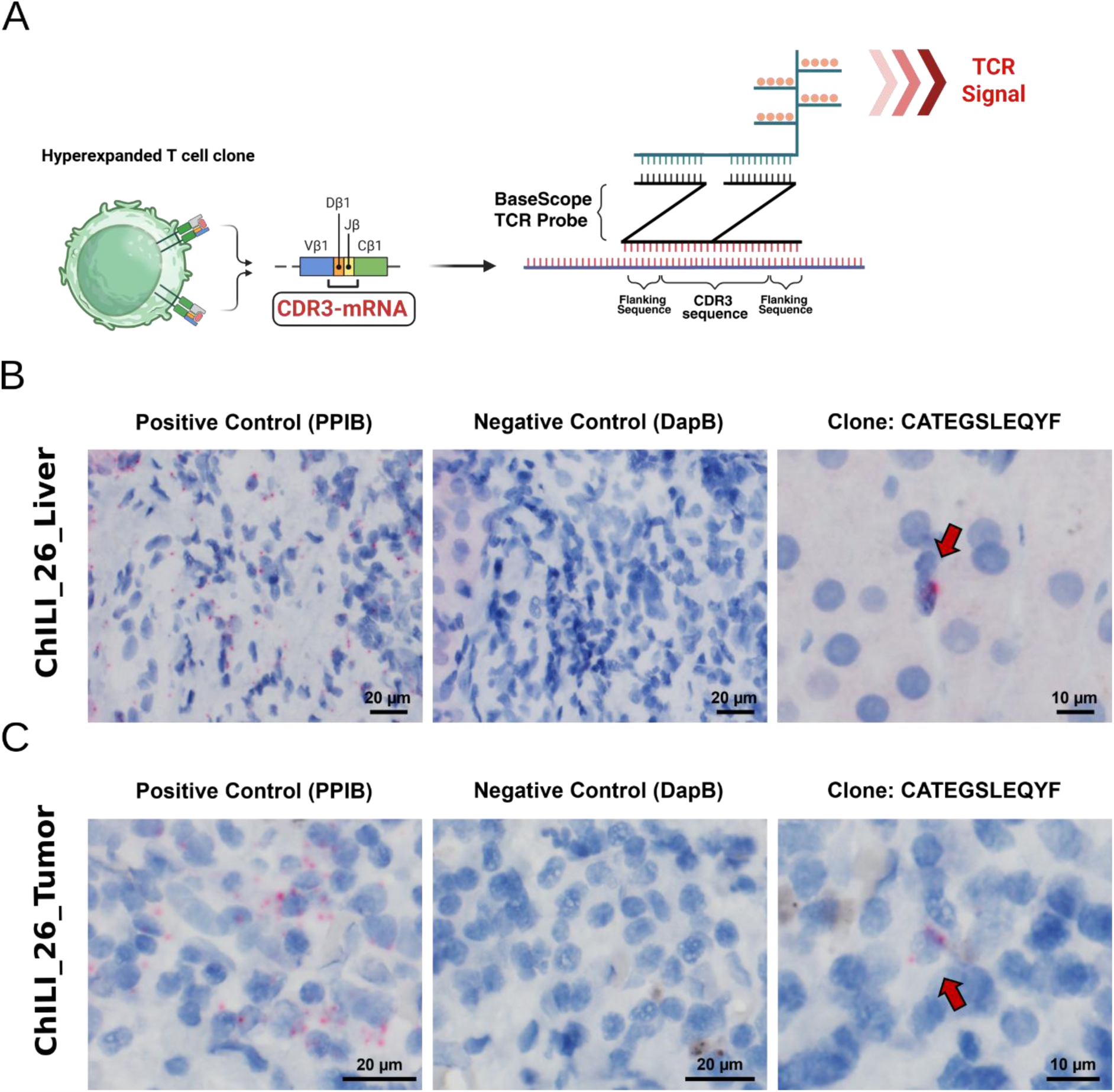
RNA in situ hybridization (RISH) with BaseScope identifies the shared hyperexpanded clones in situ. (A) A BaseScope RISH probe designed to target CDR3 and flanking mRNA nucleotide sequence of the selected T cell clone. (B) BaseScope RISH on liver sample from patient ChILI_26. A probe targeting Peptidyl-prolyl cis-trans isomerase B (PPIB) was used as a positive control and a probe targeting the bacterial gene Dihydrodipicolinate reductase (DapB) was used as a negative control. A probe (CATEGSLEQYF) was used to identify CDR3 mRNA sequence of top shared hyperexpanded T cell clones in patient ChILI_26. (C) BaseScope RISH on tumour sample from patient ChILI_26. A probe targeting Peptidyl-prolyl cis-trans isomerase B (PPIB) was used as a positive control and a probe targeting the bacterial gene Dihydrodipicolinate reductase (DapB) was used as a negative control. A probe (CATEGSLEQYF) was used to identify CDR3 mRNA sequence of top shared hyperexpanded T cell clones in patient ChILI_26.

**Extended Data Figure 10.**
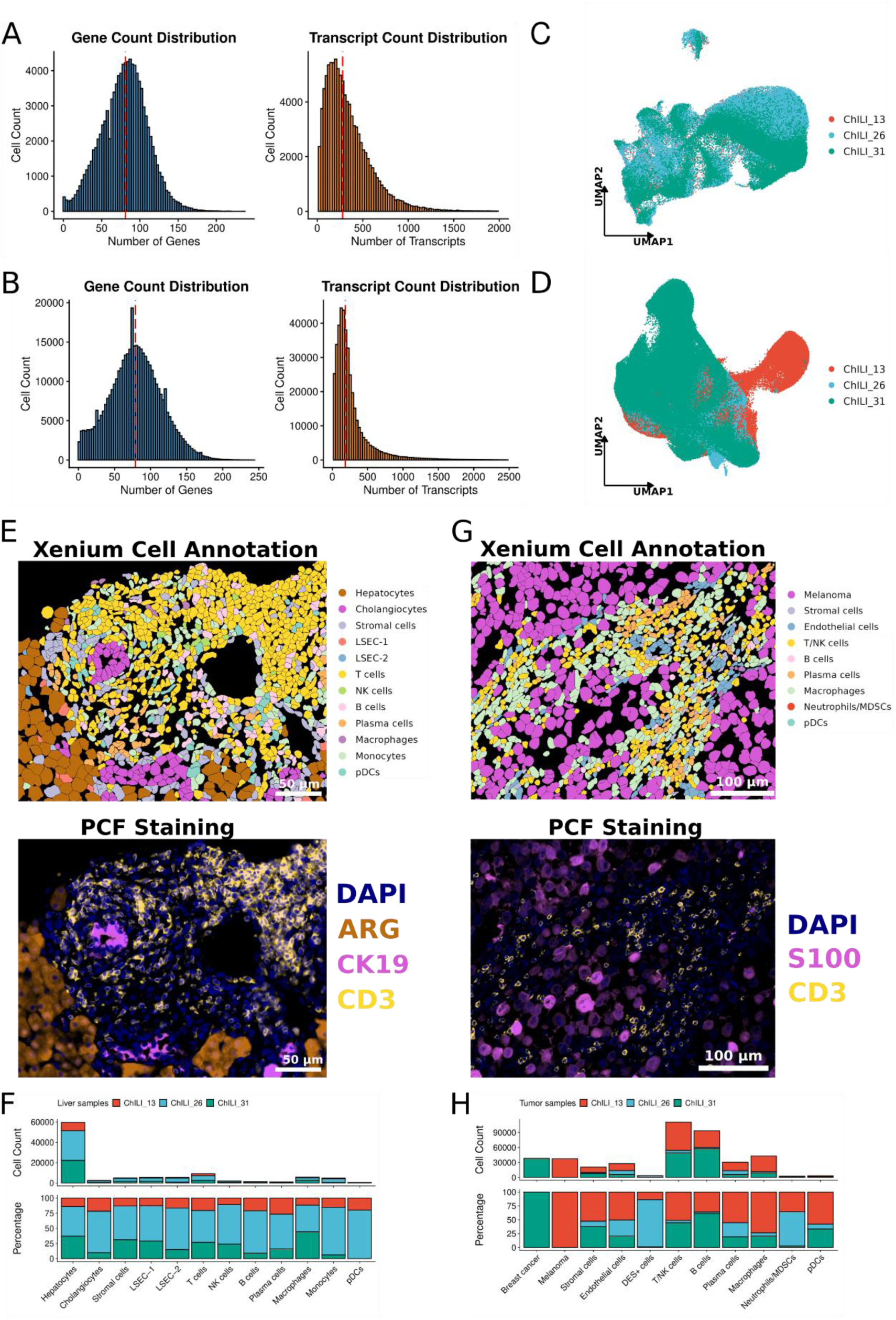
Quality control of Xenium in situ data from liver and tumour samples. (A) Gene and transcript count distribution in liver samples. (B) Gene and transcript count distribution in tumour samples. (C) UMAP projection of cells from three liver samples. (D) UMAP projection of cells from three tumour samples. (E) Top: Xenium in situ cell type annotation from a selected region in the liver sample from patient ChILI_26. Each colour shows a different cell type. Bottom: PCF staining of the same region as the upper image. (F) Count and frequency of each cell type in the liver sample. (G) Top: Xenium in situ cell type annotation from a selected region in the tumour sample from patient ChILI_13. Each colour shows a different cell type. Bottom: PCF staining of the same region as the upper image. (H) Count and frequency of each cell type in the tumour sample.

**Extended Data Figure 11.**
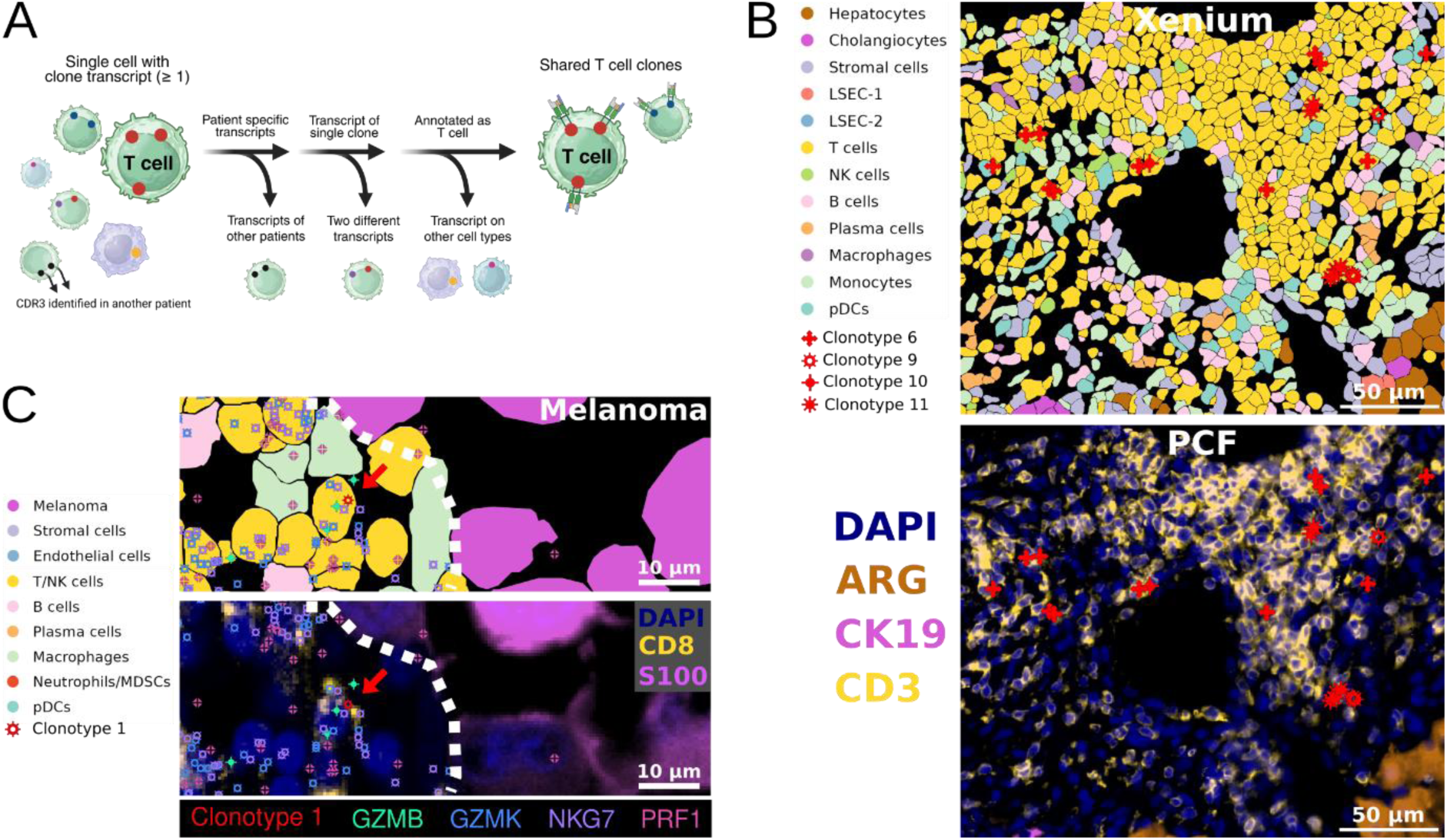
CDR3 transcripts are identified in T cells. (A) Analysis approach to identify T cells of interest with Xenium in situ. Cells with at least one clone transcript were first filtered to remove potential doublets, defined as cells containing transcripts from more than one T cell clone. Next, cells mapped to non–T cell clusters were excluded. The remaining cells were visually validated as T cells using CD3 staining from the Phenocycler-Fusion mIF data. Finally, cells that passed all these criteria were considered a single T cell clone of interest. (B) Visual validation of shared clone transcript assignment. Top: Shared clone transcripts from ChILI_26 liver sample are mapped on Xenium in situ cell type annotation. Bottom: Shared clone transcripts from ChILI_26 liver sample are mapped on PCF mIF staining of the same tissue section. Scale bar = 50 μm. (C) Visual representation of main cell clusters and shared T cell clones within the tumour of ChILI_13. Top: A region in the tumour sample showing interface between tumour and stroma. Red arrow indicates clone 1 which expresses the cytotoxicity markers *GZMB*, *GZMK*, *NKG7* and *PRF1*. Scale bar = 10 µm. Bottom: PCF mIF staining of the same region. Red arrow indicates clone 1. PCF markers were indicated as DAPI, CD8 and S100. CD8 expression of clone 1 is also confirmed at the protein level with PCF. Scale bar = 10 µm.

**Extended Data Figure 12.**
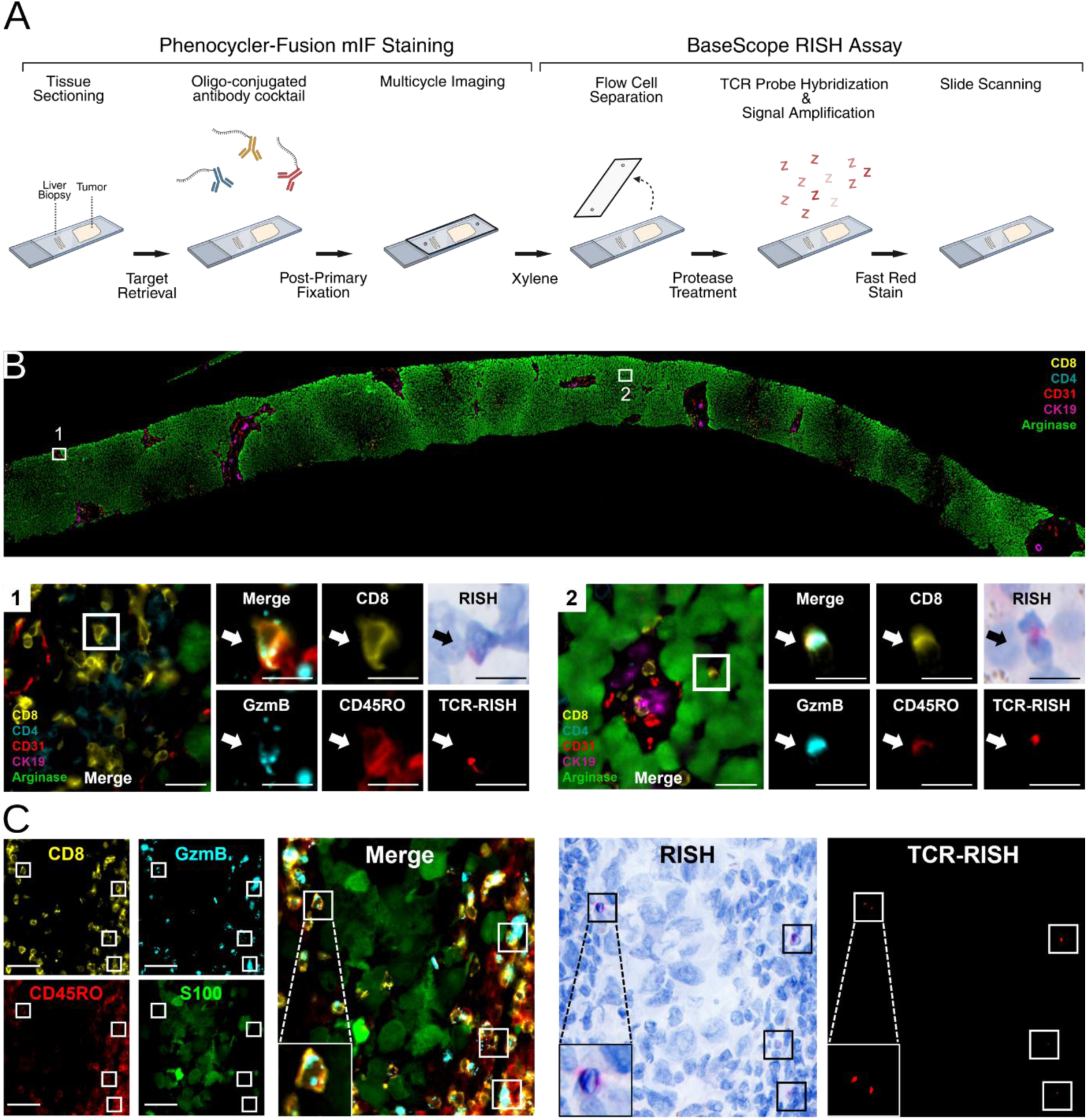
Sequential mIF and BaseScope RISH staining. (A) Workflow of sequential mIF/RISH staining. (B) 38-marker Phenocycler-Fusion staining of ChILI_13 liver biopsy (top). RISH+ cells were identified in different regions of the liver (bottom: Rectangle 1 and 2). White arrows show CASSPLTGELFF+ CD8+ GzmB+ CD45RO+ T cells. Scale bar (merge) = 20 µm, scale bar = 10 µm. (C) 38-marker Phenocycler-Fusion staining of ChILI_13 matched tumour sample. Cells inside the white rectangles are identified as CASSPLTGELFF+ CD8+ GzmB+ CD45RO+ T cells. Scale bar = 10 µm.

**Extended Data Figure 13.**
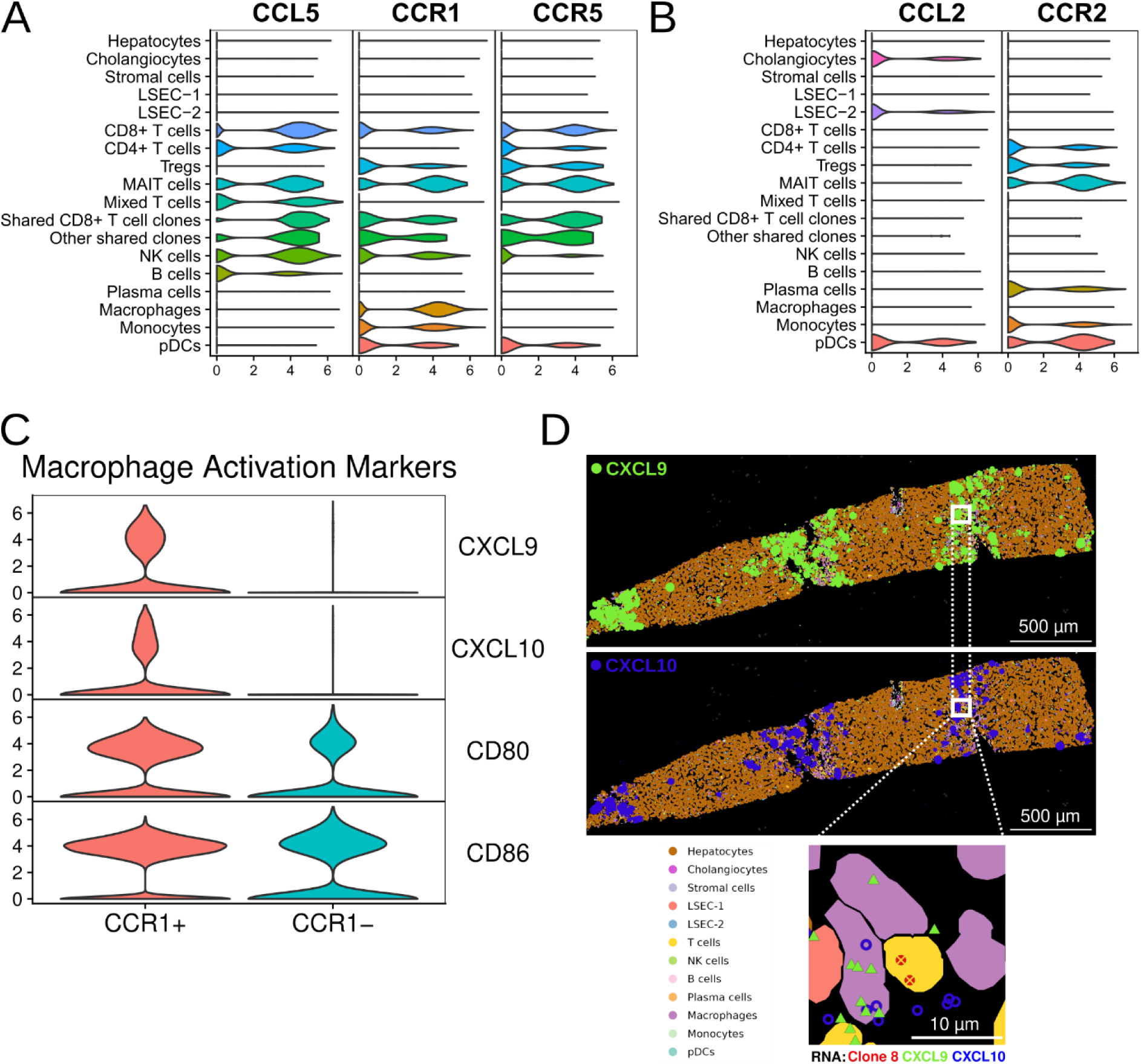
CCR1+ macrophages show activation phenotype. (A) Violin plot showing CCL5, CCR1 and CCR5 expression in different cell types. (B) Violin plot showing CCL5, CCR1 and CCR5 expression in different cell types (C) Violin plot showing expression of selected macrophage activation markers (CXCL9, CXCL10, CD80 and CD86) in CCR1+ and CCR1-macrophages. (D) Exemplary visual representation of a liver region in P31 rich in CXCL9 and CXCL10 expression. Top: Dots represent transcripts of CXCL9 (green) and CXCL10 (blue). Rectangle marks the area magnified at the bottom. Magnified area showing clone 8 and mapping of CXCL9 and CXCL10 to broad cell types annotated from spatial transcriptomics data.

**Supplementary Table 1.** Overview of ChILI cohort and normal liver cohort

Supplementary Table 1 can be found as a separate supplementary document.

**Supplementary Table 2.** Clinicopathological features of ChILI cohort

Supplementary Table 2 can be found as a separate supplementary document.

**Supplementary Table 3.** Phenocycler-Fusion antibody panel for liver biopsy TMA mIF staining

Supplementary Table 3 can be found as a separate supplementary document.

**Supplementary Table 4.** T cell Immune Repertoire

Supplementary Table 4 can be found as a separate supplementary document.

**Supplementary Table 5.** BaseScope RNA in situ hybridization probe sequences.

Supplementary Table 5 can be found as a separate supplementary document.

**Supplementary Table 6.** TCR probe list for Xenium in situ

Supplementary Table 6 can be found as a separate supplementary document.

**Supplementary Table 7.** Add-on custom gene panel for Xenium in situ

Supplementary Table 7 can be found as a separate supplementary document.

**Supplementary Table 8.** Phenocycler-Fusion antibody panel for post-Xenium mIF

Supplementary Table 8 can be found as a separate supplementary document.

**Supplementary Table 9.** PhenoCycler-Fusion antibody panel for combined mIF Staining and RISH with BaseScope Assay

Supplementary Table 9 can be found as a separate supplementary document.

